# The KD Atlas: A Multi-Omics Network Resource for Kidney Disease Research

**DOI:** 10.64898/2026.02.19.26346652

**Authors:** Yacoub A. Njipouombe Nsangou, Stefan Haug, Maria A. Ulmer, Orhan Bellur, Werner Römisch-Margl, Jürgen Dönitz, Anna Köttgen, Matthias Arnold, Gabi Kastenmüller

**Affiliations:** Institute of Computational Biology, Helmholtz Zentrum München - German Research Center for Environmental Health, Neuherberg, Germany; Department of Medical Bioinformatics, University Medical Center Göttingen, Göttingen, Germany; Institute of Epidemiology and Prevention, Faculty of Medicine and Medical Center, University of Freiburg, Freiburg, Germany; Centre for Integrative Biological Signalling Studies, Albert-Ludwigs-Universität Freiburg, Freiburg, Germany; Department of Psychiatry and Behavioral Sciences, Duke University, Durham, NC, USA; German Center for Diabetes Research (DZD), Neuherberg, Germany

## Abstract

**Background:** Kidney disease refers to a broad range of disorders that impair renal structure and function. Among these, chronic kidney disease (CKD) is the most prevalent worldwide, affecting approximately 10% of the global adult population. While large-scale omics studies have identified numerous molecular associations with kidney function and disease, these insights often remain isolated within individual data layers, hindering a systems-level understanding of the functional interplay between genes, proteins, metabolites and clinical phenotypes.

**Methods:** We developed the Kidney Disease Atlas (KD Atlas) using an extended QTL-based integration strategy combined with a composite network approach. For this purpose, we leveraged results from omics studies in population-based and kidney disease-specific cohorts from the CKDGen Consortium and other large-scale initiatives and integrated them with data from knowledge databases, inferring a comprehensive network of relationships between metabolites, proteins, genes, and kidney disease-related traits.

**Results:** We present the KD Atlas, an online resource (https://metabolomics.helmholtz-munich.de/kdatlas) integrating over 25 large studies providing disease-relevant information on 20,456 protein-coding genes, 1,962 proteins, 1,375 metabolites and 40 kidney disease phenotypes connected by more than 1.2 million relationships. Through an interactive web interface, researchers can dynamically construct context-specific molecular subnetworks and perform integrated analyses without requiring specialized bioinformatics expertise. Application showcases demonstrate the resource’s utility for providing the molecular context of KD-associated genes or metabolites and for generating novel mechanistic hypotheses.

**Conclusion:** The KD Atlas provides a global, multi-omics network view of kidney pathophysiology through an intuitive interface, empowering researchers to formulate mechanistic hypotheses and prioritize candidate targets for subsequent experimental validation.

## 1. Introduction

Kidney disease encompasses a diverse spectrum of disorders affecting renal structure and function, ranging from acute kidney injury (AKI) to chronic progressive conditions as well as glomerular diseases such as focal segmental glomerulosclerosis (FSGS) and IgA nephropathy (IgAN). While these conditions differ in etiology and clinical presentation, they often share common pathophysiological processes including inflammation, oxidative stress, fibrosis, and progressive loss of kidney function [1]. Chronic kidney disease (CKD) represents the most prevalent form of kidney disease, affecting approximately 10% of the global adult population [2]. It is a leading cause of mortality worldwide, responsible for 1.2 million annual deaths [2,3]. CKD is characterized by progressive decline in glomerular filtration rate (GFR), often accompanied by proteinuria, structural kidney damage, and systemic complications [2]. Despite advances in understanding CKD pathophysiology, effective disease-modifying therapies remain limited, and the molecular mechanisms underlying disease progression are incompletely understood [4,5].

CKD is a complex disorder influenced by genetic susceptibility, environmental factors, and comorbid conditions such as diabetes and hypertension [6]. Its pathophysiology manifests across multiple molecular layers: genetic variants influence disease risk [7, 8], transcriptional and proteomic changes reflect altered renal cellular states [9], and metabolic dysregulation results from and further exacerbates kidney dysfunction [10, 11]. This complexity necessitates a systems-level perspective, where CKD is understood not as a dysfunction of isolated genes or processes, but as a systemic perturbation of interconnected biological pathways [12].

Substantial progress has been made in cataloging molecular changes in CKD and related kidney disease. In particular, large-scale genetic consortia such as CKDGen [7] have identified hundreds of loci associated with kidney function traits through genome-wide association studies (GWAS) in over one million individuals, providing fundamental insights into the genetic architecture of CKD [7,13,14,15]. Concurrently, metabolome-wide association studies (MWAS) have revealed metabolic signatures associated with CKD progression and adverse outcomes [16, 17], while transcriptomic and proteomic profiling of kidney tissue has identified pathways central to disease pathogenesis [9, 18]. Pioneering integrative work has progressively connected genetic variants to cell-type-specific regulatory mechanisms through compartment-resolved expression quantitative trait loci (eQTL), methylation QTL (meQTL) and single-cell epigenomic data, linking GWAS signals to specific kidney cell types and disease-relevant molecular pathways [19, 20, 21]. Single-cell transcriptomics has further identified novel pathogenic cell states and immune-stromal interactions driving fibrosis [22, 23]. Most recently, a multi-ancestry GWAS in 2.2 million individuals, combined with a 32-data-type “Kidney Disease Genetic Scorecard”, integrating allele-specific expression, single-nucleus chromatin accessibility and multi-ome-based regulatory circuits, established the most comprehensive genetic blueprint of kidney function to date [24]. However, a critical gap remains: current integrative efforts have largely operated within the genetic-epigenetic-transcriptomic axis, typically not considering the proteome or metabolome. Moreover, mostly static results of the integration are provided, hindering flexible adaptation of the integration task to specific research questions.

The kidney research community has developed valuable omics resources, such as transcriptomic databases (e.g., Nephroseq), proteomic catalogs (e.g., HKUPP), multi-omics repositories (e.g., KUPKB, CKDdb) [25], including open access compartment-specific and cell-type-interaction eQTL atlases, methylation QTL, protein QTL, single-nucleus ATAC-seq, single-cell transcriptomics and spatial transcriptomics data [19, 20, 21, 24] and GWAS-focused gene prioritization tools (e.g., KidneyGPS [26]). Many of these resources enable querying and analysis of the provided data but often remain confined to single-layer interrogation rather than enabling dynamic, cross-layer integration. Network medicine provides a powerful paradigm to bridge this gap, representing biological systems as interconnected networks that can integrate diverse data types [27,28, 29]. Such multi-omics networks have proven highly successful in other complex diseases, facilitating the identification of disease modules, prioritization of therapeutic targets and generation of novel mechanistic hypotheses. For example, Menche et al. [29] integrated protein-protein interactions with disease-gene associations to construct disease modules in the human interactome. This approach successfully predicted disease-disease relationships even for phenotypically distinct conditions that share no common genes, such as asthma and celiac disease, whose network overlap revealed a shared immune pathway and was validated by high clinical comorbidity. Similarly, Cheng et al. [30] integrated drug-target interaction data with disease-associated genes in the human protein-protein interaction network to construct a comprehensive drug-disease proximity map. Using network proximity measures, they identified and validated hydroxychloroquine as protective for coronary artery disease in over 220 million patients, with the mechanism confirmed through *in vitro* experiments. While these pioneering studies demonstrate that integrated multi-omics networks can deliver meaningful biological insights and enable powerful computational predictions, the networks themselves and the analytical approaches used to interrogate them remain accessible only to computational experts. A critical unmet need persists: providing researchers without specialized bioinformatics expertise with the ability to dynamically explore integrated multi-omics networks and formulate context-specific hypotheses tailored to their own research questions.

We present the Kidney Disease Atlas (KD Atlas), a comprehensive network-based resource that provides a global, multi-omics view of kidney function and disease. Building upon a previously proposed multi-omics integration framework, which uses an extended QTL-based integration strategy combined with a composite network approach [31, 32], the KD Atlas integrates data from over 25 studies. Knowledge databases and results from large-scale omics screens in population-based cohorts represent the disease-agnostic foundation which is systematically complemented with CKD-relevant datasets and additional data types specific to renal physiology. The integrated data form a comprehensive heterogeneous network, stored using the graph database Neo4j, encompassing over 20,000 protein-coding genes, approximately 2,000 proteins, around 1,400 metabolites and 40 kidney disease-related traits connected by more than 1.2 million relationships representing statistical associations from large-scale quantitative studies. A publicly available web-based user interface (https://metabolomics.helmholtz-munich.de/kdatlas) enables researchers to dynamically construct, expand and explore context-specific molecular subnetworks surrounding genes, metabolites and kidney disease phenotypes, making the integrated network accessible without requiring specialized bioinformatics expertise. We demonstrate the utility of this resource through applications ranging from proof-of-concept validation of known biology to exploratory hypothesis generation.

## 2. Methods

To establish the KD Atlas we built upon the established technical multi-omics integration framework of the Alzheimer’s Disease (AD) Atlas [31], which provides the core methodology for data integration, storage and abstraction. The KD Atlas adapts and extends this methodology to integrate kidney-specific genomic, transcriptomic, proteomic and metabolomic data. The following sections detail the core methods, with an emphasis on kidney disease-specific data sources and adaptations. Foundational technical details of the integration framework have been described previously [31]; further details of the kidney disease-specific implementations and adaptations, including a comprehensive overview of integrated datasets, are presented in the **Supplementary Material**.

### 2.1 Overall Data Integration Strategy

The applied multi-omics integration framework relies on an extended quantitative trait locus (QTL)-based integration strategy paired with a composite network approach [31,32]. First, pairwise intra- and inter-omics relationships are extracted from individual studies (e.g., statistically significant SNP-metabolite associations from an mGWAS, or significant SNP-gene eQTL associations from GTEx) and subsequently combined into a unified network by overlaying these associations through shared entities (e.g., a SNP appearing in both an mGWAS and an eQTL study, identified by the same rsID). In the constructed heterogenous network, an edge represents a single statistically significant association (e.g., “rs715-glycine”) from a specific study, annotated with the underlying summary statistics. Both disease-independent associations between (e.g., SNP-metabolite from a metabolomics GWAS) and within (e.g., transcript-transcript from a gene co-expression analysis) different omics layers as well as disease-specific associations (e.g., SNP-trait from a GWAS) were collected from large studies. In a second step, the resulting comprehensive network of full granularity is simplified by summarizing and collapsing edges and nodes into an abstracted view with only four node types (“gene”, “metabolite”, “trait”, “meta-trait”) connected by specific edge types (e.g., “Associated in GWAS”), which then bundle same associations from multiple studies.

For the construction of the KD Atlas, we retained the core infrastructure of the AD Atlas, including the data model, which was designed to be generalizable across complex diseases, the integration pipelines, and the abstraction layers. We extended the existing framework by adding new disease-independent association data sets and by integrating protein-protein interactions as a new edge type. Finally, we systematically replaced AD-specific associations with CKD-relevant datasets and additional data types specific to renal physiology (**Supplementary Figure S1**). The two phases of the KD Atlas construction, encompassing data collection, preprocessing, formatting and integration into a comprehensive network preserving maximum granularity (Phase 1), and abstraction of this complex network into an intuitive view optimized for user interaction (Phase 2) are detailed in the following sections.

### 2.2 Phase 1: Data Collection, Preprocessing, and Integration

For the KD Atlas, we initialized the network with the set of knowledge-based relationships from public databases and non-disease-specific population-scale data that formed the core of the AD Atlas [31], including gene-transcript-protein mappings from Ensembl, SNP annotations from SNiPA, tissue-specific eQTL data from the Genotype-Tissue Expression (GTEx) Project (49 tissues), genetic associations with metabolic traits (mGWAS) from five large population-based studies, and partial correlation networks for metabolites and proteins derived from Gaussian graphical models. Together, these datasets establish a disease-agnostic molecular backbone connecting genes, metabolites and their genetic regulators across tissues, providing the generalizable framework upon which disease-specific associations are layered. Next, we extended this foundation by incorporating experimentally validated protein-protein interactions (PPI) and protein co-abundance networks from the UK Biobank, which together enrich the network with direct physical and functional relationships between proteins. Finally, we added kidney disease-specific datasets to anchor the molecular backbone in the context of kidney disease, enabling the identification of disease-relevant genes, metabolites and pathways. The integrated data were stored using the graph database management system Neo4j (version 4.4.3) [33], which natively represents network structures and enables efficient querying of complex relationships.

#### 2.2.1 Knowledge Databases and Gene Annotation

Gene-transcript-protein mappings were retrieved from Ensembl (version 97) [34], which also served as the source for primary gene, transcript and protein identifiers. Gene symbols were annotated using the HUGO Gene Nomenclature Committee nomenclature [35]. Single nucleotide polymorphism (SNP) annotations were obtained from SNiPA (version 3.3) [36]. To extend the knowledge base, experimentally validated protein-protein interactions were integrated from STRING (version 11.0) [37], HIPPIE (version 2.0) [38] and IID (version 2021.05) [39]. Interactions were restricted to experimental evidence and stratified by confidence levels (high, medium, and low) based on recurrence across databases. Detailed preprocessing procedures are provided in the **Supplementary Material**.

#### 2.2.2 Population-Based Omics Data

The following population-based datasets were included in the KD Atlas: tissue-specific eQTL data from the GTEx Project version 8 (49 tissues, including kidney cortex but not kidney medulla) [40]; genetic associations with metabolites from five large population-based studies [41–45] and partial correlation networks for metabolites and proteins derived from Gaussian graphical models [46,47]. To expand population-based metabolomic and proteomic coverage, two additional datasets were integrated: Plasma-urine metabolomics GWAS data from the German Chronic Kidney Disease (GCKD) study [48] were incorporated to capture kidney-specific metabolic processes. Protein co-abundance networks from the UK Biobank [49] were integrated to reveal additional protein-protein relationships. These datasets and their preprocessing are described in detail in the **Supplementary Material**.

#### 2.2.3 Kidney-Disease-Specific Association Data

To provide disease-focused context, kidney disease-specific association data were integrated. GWAS summary statistics for kidney function and disease traits were obtained primarily from the CKDGen Consortium [7], encompassing estimated glomerular filtration rate based on creatinine (eGFRcrea) and cystatin C (eGFRcys), urinary albumin-to-creatinine ratio (UACR), CKD, rapid kidney function decline and serum urate. Where available, diabetes-stratified analyses were included. Metabolome-wide association studies (MWAS) of kidney function and longitudinal outcomes, including kidney failure and AKI, were also integrated (full list of traits in **Supplementary Table 3**). Additionally, transcriptomic and proteomic datasets from kidney tissue, blood and plasma, as well as co-expression networks derived from RNA sequencing data, were incorporated. Weighted gene co-expression network analysis (WGCNA) [50] was applied to derive modules of positively correlated genes. Full details on the included studies, including cohort characteristics and sample sizes, are provided in **Supplementary Material**.

#### 2.2.4 Data Formatting and Integration

Each study was converted into standardized comma-separated values (CSV) files with one biological relationship per row (e.g., variant, gene, statistical measures) accompanied by metadata files documenting provenance (publication details, cohort information, sample characteristics). Integration across studies was achieved by overlaying knowledge-based and data-driven relationships. Statistical associations were represented through intermediate nodes containing summary statistics, with each result linked to its source to maintain detailed provenance.

#### 2.2.5 Biological Entity Identifier Mapping

To enable integration through entity overlap, biological entities were mapped to unique identifiers. For SNPs, rsID; and for genes, transcripts and proteins, Ensembl IDs were used as primary identifiers. For metabolites, we retained platform-specific identifiers for uniqueness. Supplementary identifiers including gene symbols (HGNC nomenclature), biochemical metabolite names and UniProt accessions were annotated but not required to be unique. Kidney disease phenotypes from different studies were harmonized through manual curation and are comprehensively listed in **Supplementary Tables 2** and **3**.

### 2.3 Phase 2: Network Abstraction and Simplification

The highly granular network resulting from Phase 1 was projected into a simplified representation to enhance interpretability and enable intuitive visualization as previously described [31]. Briefly, the abstracted view consists of four primary node types: protein-coding genes, metabolites, traits (kidney disease phenotypes) and meta-traits (collections of related phenotypes) derived through the following projections:

#### 2.3.1 Gene-Level Projection

SNPs, transcripts and proteins were mapped to their corresponding genes using multiple information sources. Proteins were linked to genes via transcript mappings from Ensembl. SNP-to-gene assignments utilized: (i) genomic location - variants within gene bodies or 2.5kb flanking regions, and variants in gene-associated regulatory elements (ENCODE promoters and enhancers [51], FANTOM5 enhancers [51]); and (ii) functional evidence - significant eQTL and pQTL associations from GTEx [34] across all 49 tissues. This approach enables one-to-many SNP-to-gene mappings, ensuring GWAS signals capture the regulatory complexity of each locus. Through the gene-level projection in the abstraction phase, the resulting gene nodes aggregate information across omics layers (transcriptomics, proteomics), i.e., an edge representing a significant co-expression between transcripts of two genes and significant co-abundance of the proteins encoded by the same two genes, are represented by two edges connecting these two genes. Only protein-coding genes were retained in the current version.

#### 2.3.2 Cross-Platform Metabolite Integration

Metabolites measured across multiple platforms were consolidated into unified "meta-metabolite" nodes through manual curation of platform-specific identifiers. For the Metabolon platform, metabolites sharing identical biochemical names and chemical identifiers but differing compound IDs were merged. Consolidation statistics are provided in **Supplementary Table S5**.

#### 2.3.3 Association Filtering and Threshold Application

Statistical associations of the same type between the same entities coming from multiple studies were summarized while preserving underlying variant-level and protein-level information in comprehensive edge annotations. Edges were filtered using study-specific, gene-wise, or genome-wide significance thresholds. For genetic associations with kidney disease traits (trait-QTLs) and metabolites (mQTLs), users can select either genome-wide significance (p ≤ 5×10⁻⁸) or gene-wise thresholds via the web interface. Gene-wise significance was defined as p ≤ 0.05/n, where n represents the number of SNPs annotated to each gene. Study-specific thresholds used for all other association types are listed in **Supplementary Table S6**. This abstraction preserves essential biological relationships while substantially reducing network complexity. More details on the abstraction procedures are provided in **Supplementary Material**.

### 2.4 Web Interface and Technical Implementation

The KD Atlas user interface was implemented by building on the AD Atlas web framework [31]. The application was developed in R (version 4.3.1) using Shiny and deployed via ShinyProxy. Communication between the Neo4j (version 4.4.3) [33] graph database and R was established through the official Neo4j Python driver using the reticulate (version 1.34) R-Python interface. Interactive network visualizations were generated with VisNetwork, employing layouts computed by igraph and qgraph algorithms. Gene enrichment analyses were performed using topGO (version 2.54.0) [52], enrichR (version 3.2) [53] and gprofiler2 (version 0.2.2) [54]. Metabolite enrichment was conducted using platform-specific annotations of metabolites into classes and super-/sub-pathways where available. Enrichment was assessed using Fisher’s exact tests (R function fisher.test, alternative = “greater”) and the resulting p-values were adjusted for multiple comparisons using the false discovery rate method (p.adjust, method = “fdr”). Network rendering options include 2D and 3D layouts, hive plots and interactive bar/donut plots, enabling users to explore nodes and edges to inspect attributes and metadata. Further implementation details are provided in the **Supplementary Material**.

## 3. Results

In this study, we created the KD Atlas, a comprehensive, network-based catalogue of multi-omics data for kidney disease, accessible through an interactive web interface at https://metabolomics.helmholtz-munich.de/kdatlas (**Figure 1**). Building upon the integration framework established for AD Atlas [31], this resource integrates data from over 25 studies, including knowledge databases, population-based omics datasets and kidney disease-specific association studies, into a large heterogeneous network capturing all entities and their relationships. To enhance accessibility and interpretability, the integrated data were abstracted into a simplified network representation consisting of four primary node types: genes (including associated transcripts, SNPs and proteins), metabolites (mapped across available platforms), traits (kidney disease phenotypes) and meta-traits (collections of related traits). Within this simplified representation, researchers can dynamically construct molecular subnetworks contextualizing entities of interest, including genes, metabolites or kidney disease phenotypes (**Figure 2**). Built-in analysis tools allow to interactively conduct downstream analyses of these context-specific subnetworks without the need for advanced bioinformatics skills. In three showcases, we exemplify the applicability of our resource for validating established kidney disease mechanisms (*UMOD* tubular transport network, *CUBN* receptor complex) and generating novel hypotheses (gut microbiome-derived metabolites linking to glomerular disease).

**Figure 1.**
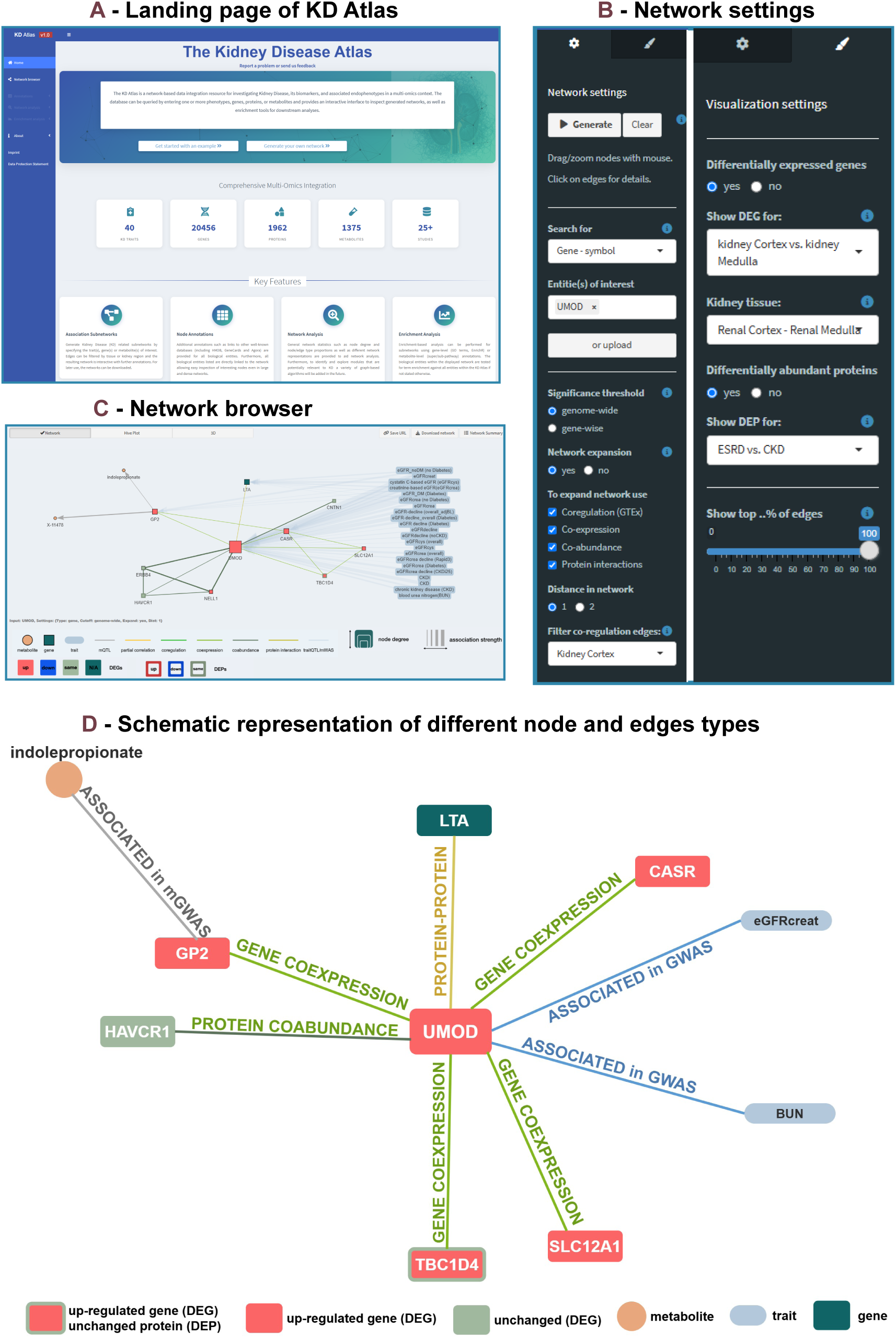
Overview of the KD Atlas web interface and network visualization features. **A**. Landing page presenting summary statistics and entry points to network generation modules. **B**. Configuration panels for defining parameters to generate molecular subnetworks (left) and for overlaying differential expression data for genes (DEG) and proteins (DEP) (right). **C**. Network browser displaying a multi-omics subnetwork centered on *UMOD*, where nodes represent genes, metabolites and kidney disease traits connected through molecular relationships. Node colors indicate differential expression status when DEG data are overlaid: upregulated genes (red), downregulated genes (blue), and genes without significant differential expression (muted sage green). **D**. Schematic representation of the same *UMOD*-centered subnetwork shown in C, illustrating node types and a subset of the relationships included in this example. For simplicity, only selected nodes and edges are shown. Abbreviations: Genome-wide association study (GWAS); metabolome-wide GWAS (mGWAS)

**Figure 2.**
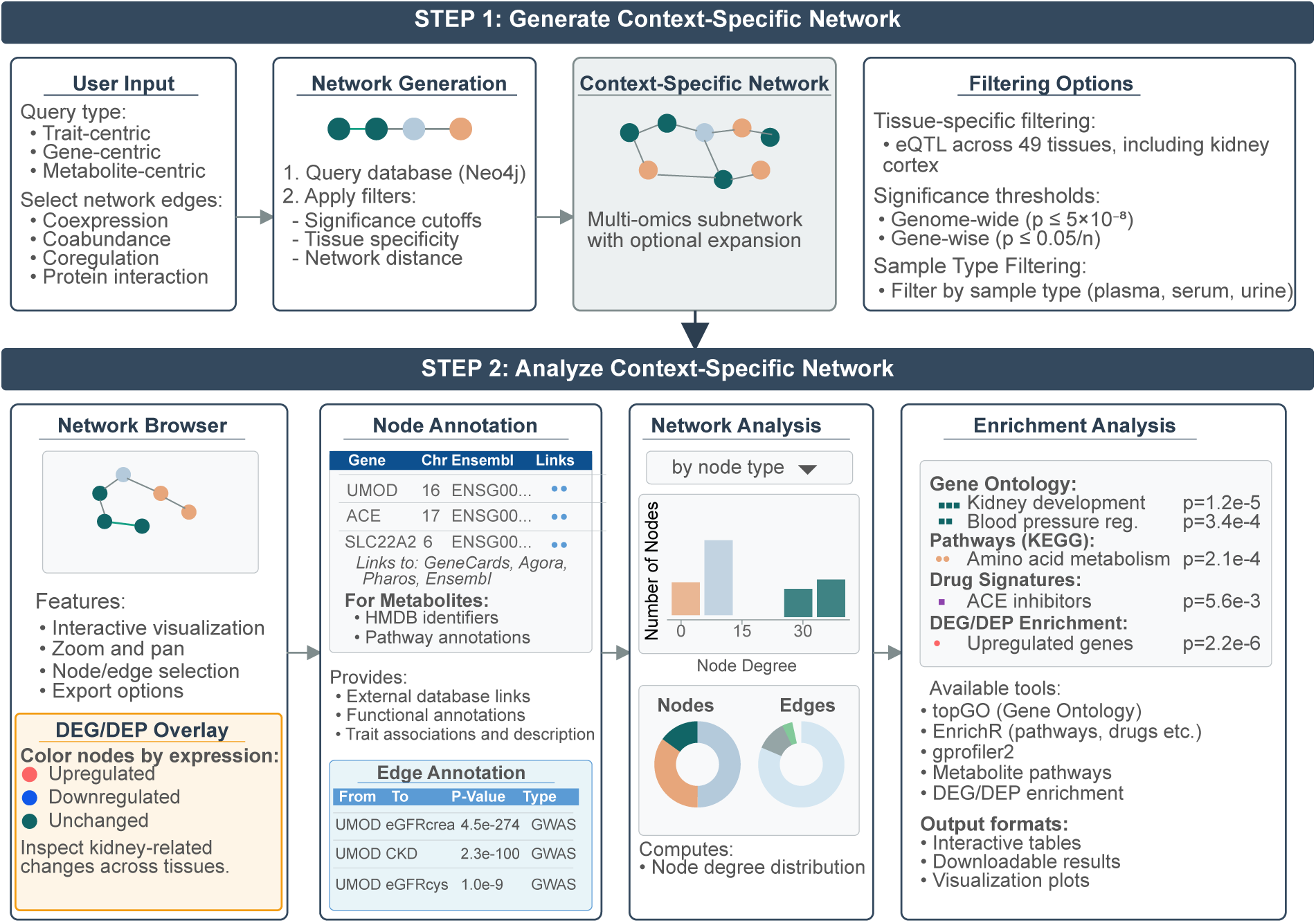
Interactive workflow for generating and analyzing context-specific networks in the KD Atlas. Step 1: Users generate customized molecular subnetworks through trait-centric, gene-centric or metabolite-centric queries with optional network expansion and filtering options including tissue-specific eQTL selection across 49 tissues (including kidney cortex), significance thresholds (genome-wide p ≤ 5×10⁻⁸ or gene-wise p ≤ 0.05/n, where n is the number of SNPs annotated to the gene) and sample type filtering. Step 2: Generated networks support interactive exploration, differentially expressed gene/differentially expressed protein (DEG/DEP) overlay to visualize kidney-related expression changes, analysis across multiple databases including Gene Ontology, KEGG pathways, drug signatures, metabolic pathways and DEG/DEP enrichment.

### 3.1 The KD Atlas Content

The KD Atlas comprises 20,456 protein-coding genes, 1,962 proteins, 1,375 metabolites, 40 kidney disease traits and 6 meta-traits (**Table 1**). Kidney disease traits include kidney function measures such as eGFRcrea, eGFRcys, blood urea nitrogen (BUN), UACR and clinical outcomes including CKD incidence, rapid kidney function decline and kidney failure (**Supplementary Table 3**). Both cross-sectional measures and longitudinal outcomes are included, with diabetes-stratified analyses incorporated where available to capture gene-by-diabetes interactions.

**Table 1.** Overview over data integrated within the KD Atlas (simplified data view).

| Nodes | Count (n) | Notes |
| --- | --- | --- |
| Meta-traits | 6 | Collections of related kidney disease traits (e.g., 'eGFR' grouping eGFRcrea, eGFRcys and eGFR decline measures; 'CKD' grouping CKD incidence and progression traits) |
| Traits | 40 | CKD and kidney related phenotypes (eGFR, UACR, CKD incidence/progression, etc.) |
| Gene (protein-coding) | 20,456 | Includes associated transcripts, SNPs, and proteins |
| With DEG information | 18,983 | Covers DEGs from $\geq 1$ kidney-related phenotypes [55-57, 59] |
| With DEP information | 1,966 | Covers DEPs from $\geq 1$ kidney-related phenotypes [58] |
| Metabolites | 1,375 | Mapped across available platforms |
| **Relationships** |  |  |
| Genetic associations with kidney disease traits (trait-QTLs) | 34,193 | SNP-trait associations from CKDGen Consortium and related studies [13-15, 60-67] |
| Metabolite associations with kidney disease traits (MWAS) | 121 | Metabolite-trait associations from MWAS studies [16-17, 68-69] |
| Genetic associations with metabolites (mQTLs) | 98,253 | SNP-metabolite associations [41-45, 48] |
| Gene co-regulation (tissue-specific eQTLs, including 6,996 from kidney cortex) | 4,008,088 | Across 49 tissues [40] |
| Gene co-expression | 600,562 | Includes significant positive gene-gene correlations (derived using WGCNA-based networks)* |
| Protein co-abundance | 32,406 | Derived through partial correlation of plasma protein abundances [47, 49] |
| Metabolite co-abundance (data-derived metabolic pathways) | 794 | Derived through partial correlation of serum metabolite abundances [46] |
| Protein-protein interaction | 334,416 | Includes experimentally validated PPIs from STRING, HIPPI and IID (high-confidence: 107,924, medium: 165,118, low: 61,374) [37-39] |

\* For additional methodological details on gene co-expression network construction, see **Supplementary Material**, Section 2.2.6.

Biological entities and traits are interconnected by over 1.2 million relationships representing statistical associations inferred from large-scale quantitative studies (**Table 1**). These relationships include: 34,193 genetic associations with kidney disease traits (trait-QTLs); 98,253 genetic associations with metabolic traits (mQTLs); 121 metabolite associations with kidney disease traits identified through MWAS; 600,562 gene co-expression relationships in kidney tissue; 4,008,088 tissue-specific eQTL associations from 49 tissues (with 6,996 specific to kidney cortex); 32,406 protein co-abundance and 794 metabolite co-abundance edges derived through partial correlation analysis; and 334,416 experimentally validated protein-protein interactions classified by confidence level (107,924 high-confidence, 165,118 medium-confidence and 61,374 low-confidence). **Figure 1D** depicts examples of these relationship types taken from the subnetwork surrounding *UMOD* (**Figure 1C**).

In addition, results from differential gene expression and protein abundance studies can be projected on the network nodes, enabling downstream enrichment analyses within the dynamically constructed subnetworks (see **Supplementary Material**, Sections 2.2.4–2.2.5 for details on kidney compartments, disease contexts and comparisons). For these projections, results for, in total, 18,983 genes and 1,966 plasma proteins are available from multiple studies, including glomerular comparisons (FSGS vs. minimal change disease vs. controls) [55], kidney compartment comparisons (cortex vs. medulla) [56], disease-specific analyses (IgA nephropathy vs. healthy donors) [57], and CKD progression (early vs. end-stage CKD) [58].

### 3.2 User Interface and Network Exploration

The KD Atlas web interface provides three main entry points – trait-centric, gene-centric and metabolite-centric – for building context-specific molecular subnetworks (**Figure 2**). Users define their entities of interest. Additionally, they can customize the subnetwork construction by controlling the applied significance thresholds (genome-wide or gene-wise), filtering by tissue or sample types, and choosing the depth of network expansion (**Figure 1B**, see **Supplementary Material** Section 5 for details).

- *Trait-centric subnetworks:* Users select one or more kidney disease traits or meta-traits. The system retrieves associated genes and metabolites (via direct trait-QTL and MWAS edges), adds additional genes and metabolites that are linked via mQTL edges, and integrates their relationships (i.e., filling in all edges that directly connect any of these entities) while considering the customization options (e.g., only using co-expression etc. from a specific tissue).
- *Gene-centric subnetworks:* Users provide one or more genes identified by HGNC symbol or Ensembl ID. The system retrieves metabolites (via mQTL) and traits (via trait-GWAS) directly associated with the specified genes and includes relationships among all extracted entities according to the selected options. The initial gene set can be expanded to include their 1- or 2-step neighbors based on four types of molecular associations: gene co-expression, protein co-abundance, genetic co-regulation (eQTL data) and experimentally validated protein–protein interactions. Tissue-specific filtering can be applied to restrict expansion to kidney-relevant functional neighborhoods.
- *Metabolite-centric subnetworks:* Users provide one or more metabolites by biochemical name or query by metabolic pathway categories (SuperPathway, SubPathway) or chemical class. The system extracts traits (via MWAS) and genes (via mQTL) directly associated with the specified metabolites and includes all relationships among the retrieved entities according to the selected options. The their partial correlations with further metabolites (i.e., metabolite co-abundance edges).

The generated networks can be explored visually via the network browser (**Figure 1C**) and overlaid with results from analyses of differential gene expression and/or protein co-abundance, allowing transcriptional and proteomic changes between various groups (e.g., cortex vs. medulla, FSGS vs. control, early vs. late-stage CKD) and strata (e.g., with/without diabetes) to be examined across relevant tissue contexts (**Figure 1C**). Up- and downregulation of genes and proteins is indicated by node and border coloring, respectively, with further visualization options described in **Supplementary Material,** Section 5.

### 3.3 Network Analysis and Enrichment Tools

Context-specific subnetworks generated in the KD Atlas are accompanied by network statistics computed on the fly, including node degree distributions and entity compositions. Moreover, we provide tools for gene set and pathway enrichment analyses, allowing the entities in the network to be functionally characterized. Users can export networks and analysis results in multiple formats for downstream use (**Figure 2**). Detailed information on interactive visualization, subnetwork annotation, enrichment analyses and data export options is provided in **Supplementary Material,** Section 5.

### 3.4 Application Showcases Demonstrating the Utility of Context-Specific Network Analysis

To demonstrate the utility of the KD Atlas, we present three examples for its application. We organize these applications into two categories: (1) proof-of-concept showcases, demonstrating that the KD Atlas accurately captures well-characterized kidney disease biology, and (2) hypothesis-generating showcases, to discover potential novel molecular connections that warrant experimental validation. For proof-of-concept showcases (Sections 3.4.1–3.4.2), we focus on genes with established roles in kidney physiology such as *UMOD* and *CUBN* to demonstrate that the network-based integration correctly reconstructs known protein complexes and functional pathways. For the third, hypothesis-generating showcase (Section 3.4.3), we adopt an exploratory approach starting from disease-associated gut microbiome-related metabolites to possibly identify yet unknown molecular relationships that generate testable hypotheses regarding disease mechanisms. All showcases utilize the KD Atlas web interface to construct context-specific molecular subnetworks, overlay differential expression data, and perform functional enrichment analyses.

#### 3.4.1 UMOD Tubular Transport Network Highlights Kidney-Specific Ion Homeostasis and Immune Function

Uromodulin (*UMOD*), also referred to as Tamm-Horsfall protein, is a glycoprotein synthesized exclusively by epithelial cells of the thick ascending limb of the loop of Henle and early distal tubules, and constitutes the most abundant protein found in human urine under physiological conditions [70,71]. While its full range of biological functions continues to be elucidated, established roles include regulation of tubular ion transport, immunomodulation, and protection against urinary tract infections and kidney stones [70,71]. Genetic studies have highlighted *UMOD* across a spectrum of kidney disease: among the strongest genetic determinants of CKD risk and hypertension in the general population [70,72]. We used the KD Atlas to contextualize *UMOD* within its multi-omics molecular neighborhood and investigate the biological pathways underlying these genetic associations.

We queried the KD Atlas using *UMOD* as input and expanded the network to include genes in its 1-step neighborhood based on gene co-expression, genetic coregulation (eQTL), protein co-abundance, and experimentally validated protein–protein interactions data. We applied a genome-wide significance cutoff and filtered co-regulation edges for ’Kidney Cortex’ to construct high-confidence, tissue-specific subnetworks. The resulting network consists of 10 protein-coding genes, 2 metabolites and 22 kidney disease traits (**Figure 3A**). Visual inspection of the network reveals *UMOD* as the central hub with strong connections to multiple genes through co-expression and co-abundance relationships, as well as extensive associations with kidney function traits.

**Figure 3.**
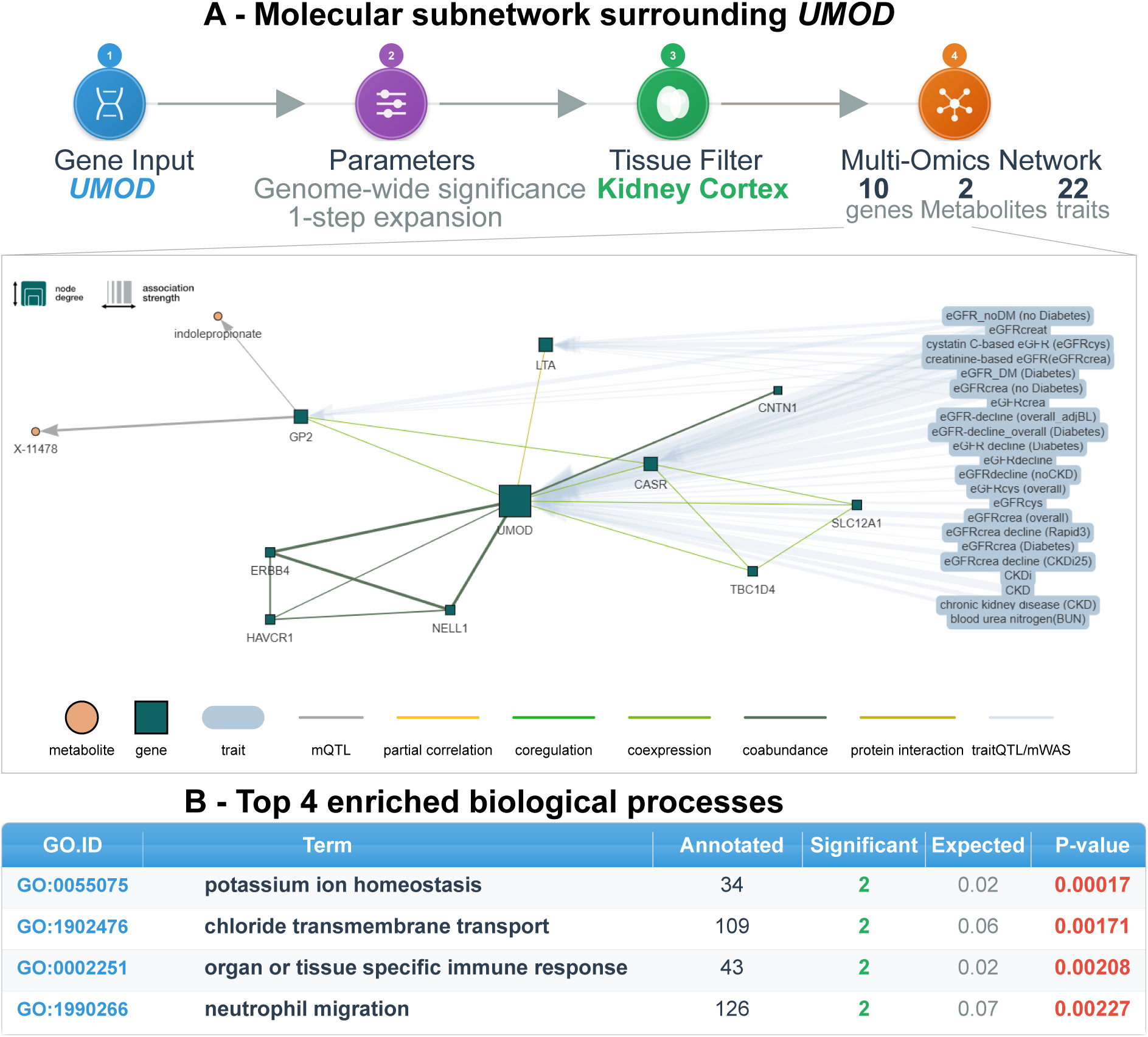
*UMOD* subnetwork highlights its link to tubular transport and immune function. **A.** Context-specific, multi-omics subnetwork surrounding *UMOD* within the KD Atlas. **B.** Gene Ontology enrichment analysis using topGO reveals significant enrichment of ion homeostasis and immune response processes.

The co-expressed genes surrounding *UMOD* include *SLC12A1*, *CASR*, *TBC1D4* and *GP2*, all of which show kidney-specific expression patterns. *SLC12A1* encodes the Na-K-2Cl cotransporter 2 (NKCC2), a key transporter in the thick ascending limb that is responsible for sodium, potassium and chloride reabsorption and represents the pharmacological target of loop diuretics [73]. *CASR* encodes the calcium-sensing receptor, which is expressed in multiple kidney segments including the thick ascending limb, where it regulates calcium reabsorption and is the target of calcimimetic drugs used in CKD [74]. *TBC1D4* encodes a Rab GTPase-activating protein involved in glucose transporter trafficking and has been implicated in metabolic regulation [75]. *GP2*, a structural paralog of *UMOD* [76], appears in the network connected to the metabolite indolepropionate (a metabolite associated with reduced risk of type 2 diabetes and inflammation [78], see also Section 3.4.3) through metabolite quantitative trait locus (mQTL) associations. The network also includes *NELL1*, *HAVCR1*, *ERBB4* and *CNTN1*, which are connected through co-abundance relationships of the encoded proteins. Notably, *LTA* (lymphotoxin alpha), a cytokine secreted by activated lymphocytes, connects to *UMOD* through a protein-protein interaction, suggesting potential links to immune regulation.

Four genes in the network (*UMOD*, *LTA*, *CASR* and *GP2*) harbor genetic variants associated with kidney disease traits. The trait connectivity includes 22 phenotypes covering the spectrum of kidney function, including eGFRcrea, eGFRcys, diabetes-stratified measures (eGFR_DM, eGFR_noDM), longitudinal and rapid decline phenotypes (Rapid3, CKDi25), CKD incidence and quantitative kidney function markers such as blood urea nitrogen (BUN). This comprehensive connectivity highlights the central role of *UMOD* and its molecular neighborhood in kidney function across multiple physiological and pathological contexts.

To functionally characterize the biological processes assigned to the genes in this subnetwork, we performed Gene Ontology (GO) enrichment analysis using the topGO [52] tool within the KD Atlas. The most significantly enriched biological processes were potassium ion homeostasis, chloride transmembrane transport, organ- or tissue-specific immune response and neutrophil migration (see table **in Figure 3B**). Additional enriched terms included ERBB2–ERBB4 signaling pathway and ERBB4–ERBB4 signaling pathway, reflecting the presence of *ERBB4* in the co-abundance network and its role in cellular signaling. These enrichment results align remarkably well with the known functions of *UMOD* described in the literature. Devuyst et al. and LaFavers et al. comprehensively reviewed *UMOD* biology and highlighted its roles in regulating salt transport in the thick ascending limb, immunomodulation, and protection against urinary tract infections and kidney stones [70,71]. The enrichment of ion transport processes (potassium homeostasis, chloride transport) is consistent with the physiological context of *UMOD* expression in the thick ascending limb, where it is co-expressed with key ion transporters including the *SLC12A1*-encoded NKCC2 cotransporter [70]. The enrichment of immune-related processes (organ-specific immune response, neutrophil migration) is consistent with the immunomodulatory functions of *UMOD*, including its ability to modulate innate immune responses and provide protection against urinary pathogens [70,71].

The network structure provides molecular context for understanding how genetic variation at the *UMOD* locus influences kidney function. Common variants in the *UMOD* promoter that increase *UMOD* expression are associated with reduced risk of CKD and hypertension [70,71]. This observed genetic association aligns with *UMOD*’s role in regulating electrolyte transport in the thick ascending limb, where it is expressed alongside key transporters such as NKCC2 (*SLC12A1*) [77]. The co-expression relationship between *UMOD* and *SLC12A1* captured by the KD Atlas is consistent with this shared physiological context and demonstrates the ability of the network to recover biologically meaningful molecular relationships.

In conclusion, this analysis demonstrates how the KD Atlas can contextualize well-studied genes within their comprehensive multi-omics molecular neighborhood. The *UMOD* network captured biologically meaningful co-expressed genes involved in tubular ion transport (*SLC12A1*, *CASR*), highlighted these connections through GO enrichment analysis identifying the expected ion homeostasis and immune functions, and linked these molecular relationships to the extensive genetic associations between *UMOD* variants and kidney function measures across multiple cohorts. Notably, genes in this network include established pharmacological targets (*SLC12A1* for loop diuretics [73,79], *CASR* for calcimimetics [74]), highlighting the clinical relevance of these pathways. This showcase thus illustrates the utility of the KD Atlas for extracting known disease-related biology and providing mechanistic context to genetic findings in kidney disease.

#### 3.4.2 CUBN Network Reveals Multi-Ligand Receptor Complex for Albumin and Vitamin Reabsorption

Cubilin (*CUBN*) is a large multi-ligand receptor that is highly expressed in renal proximal tubule epithelial cells, where it mediates the reabsorption of filtered proteins, lipoproteins and vitamins. It is also expressed in glomerular podocytes and in the intestinal epithelium, where it contributes to vitamin B12 (cobalamin) absorption [80]. *CUBN* does not possess a transmembrane domain and therefore requires binding partners to mediate endocytosis and intracellular trafficking. The receptor functions as part of a complex with megalin (*LRP2*), another multi-ligand receptor with an extensive transmembrane domain, and amnionless (*AMN*), a single-pass transmembrane protein that anchors *CUBN* to the cell membrane [80–83]. Mutations in *CUBN* cause Imerslund-Gräsbeck syndrome, an autosomal-recessive disorder characterized by vitamin B12 (cobalamin) malabsorption leading to megaloblastic anemia, often accompanied by proteinuria [84]. Common genetic variants near the *CUBN* encoding gene have been associated with urinary albumin excretion, a well-established marker of kidney damage and risk factor for CKD, in genome-wide association studies [64, 66], suggesting broader involvement of *CUBN*-mediated pathways in kidney disease susceptibility.

We used the KD Atlas to investigate the molecular neighborhood of *CUBN* and evaluate whether the network captures known functional partnerships and biological processes. We queried the KD Atlas using *CUBN* as input gene and expanded the network to include 1-step neighbors based on co-expression, genetic co-regulation, protein co-abundance, and protein-protein interaction data. The subnetwork was constructed by filtering co-regulation edges for the tissue ‘Kidney Cortex’ and applying a genome-wide significance cutoff. The resulting network contains 57 protein-coding genes, 126 metabolites and 17 kidney disease traits (**Figure 4A**). Among the 57 genes in the network, 21 harbor genetic variants associated with kidney disease traits, including *CUBN* itself, *LRP2*, *DDC*, *HNF4A* and multiple solute carriers and metabolic enzymes. Among the 126 metabolites, 14 showed direct associations with kidney disease traits, including carnitine species (acetylcarnitine, propionylcarnitine, undecenoylcarnitine), amino acids (phenylalanine, glycine) and androsterone glucuronide, suggesting that *CUBN*-associated genetic variation may influence multiple aspects of metabolic homeostasis relevant to kidney function.

**Figure 4.**
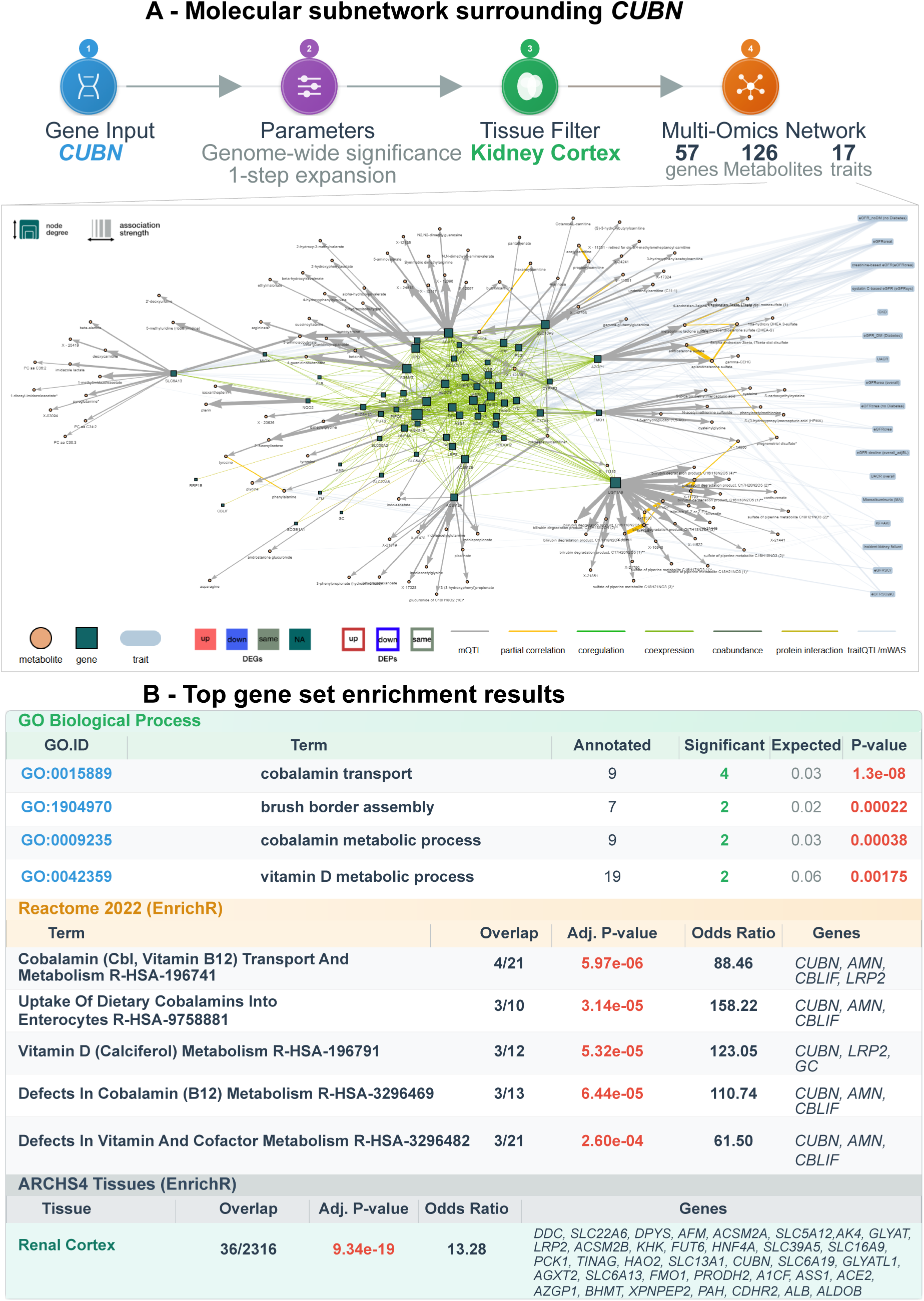
*CUBN* network reconstructs multi-ligand receptor complex and highlights vitamin reabsorption pathways. **A.** Multi-omic subnetwork surrounding *CUBN* from the KD Atlas. **B.** Enrichment analysis across multiple databases (Gene Ontology via topGO, Reactome 2022 and ARCHS4 Tissues via EnrichR) identifies cobalamin (B12) transport and metabolism as the top pathways, with extreme tissue-specific enrichment in the renal cortex.

Visual inspection of the network immediately revealed the presence of *LRP2* (megalin) and *AMN* (amnionless) among the genes connected to *CUBN*, confirming that the KD Atlas successfully captures the known protein complex required for *CUBN* function. *LRP2* showed a co-expression relationship with *CUBN*, consistent with their coordinated expression in proximal tubule cells where they jointly mediate protein reabsorption [85]. *AMN* was connected to *CUBN* through a protein–protein interaction, reflecting its role as the membrane anchor required for cubilin localization and function [85,86]. The recovery of this well-characterized receptor complex demonstrates that network-based integration of co-expression and interaction data can identify functionally related proteins that operate together in specific cellular processes. Among the 17 traits associated with the genes in the network, UACR was present, connecting to genes including *CUBN*. This genetic association reflects the role of *CUBN* in albumin reabsorption in the proximal tubule; variants that reduce *CUBN* expression or function would be expected to decrease albumin reabsorption capacity, leading to increased urinary albumin excretion [64,85–87]. The network also includes connections to multiple eGFR definitions and kidney function decline measures, suggesting that genetic variation in *CUBN* and its associated molecular neighborhood may influence broader aspects of kidney function beyond albumin handling alone.

Enrichment analysis confirmed that the *CUBN* network represents biologically coherent processes. Gene Ontology and Reactome analyses identified cobalamin (vitamin B12) transport and metabolism as the most significantly enriched terms, directly reflecting *CUBN*’s established role in intestinal vitamin B12 absorption and renal reabsorption of filtered cobalamin [84]. Vitamin D metabolism and brush border assembly were also enriched, consistent with *CUBN*’s apical localization and its function in reabsorbing vitamin D-binding protein complexes [88]. Tissue enrichment analysis identified renal cortex as the most significantly enriched tissue, with contributing genes including *CUBN* and *LRP2*, confirming kidney-specific expression of the network (see table **in Figure 4B**). Importantly, renal cortex enrichment was observed regardless of whether kidney cortex tissue was selected during network construction, confirming that this result reflects genuine biological signal rather than a filtering artifact.

Overall, the *CUBN* network captured the known *CUBN*-*LRP2*-*AMN* receptor complex, key vitamin and protein reabsorption processes and renal cortex specificity. This molecular context supports genetic findings linking *CUBN* variation to albuminuria and kidney function traits, demonstrating how KD Atlas data integration can help to extract known biology and provide systems-level insight into disease-relevant pathways.

#### 3.4.3 Gut Microbiome-Derived Metabolites Link to Glomerular Disease Through Genetic Variation in Amino Acid Metabolism and Organic Cation Transport

Gut microbiome-derived metabolites, particularly those derived from aromatic amino acid metabolism, have emerged as important contributors to uremia and CKD progression [89,90]. Tryptophan-derived indole metabolites such as indoxyl sulfate and p-cresol sulfate are well-established uremic toxins that accumulate in CKD patients and have been associated with cardiovascular complications, inflammation and kidney disease progression [91,92]. However, the genetic architecture linking gut microbial metabolism to kidney disease risk remains incompletely characterized. Traditional MWAS can identify direct metabolite-disease associations, but they may miss indirect connections mediated through genetic regulation of metabolite-processing enzymes and transporters. We hypothesized that leveraging the integrated mQTL and kidney function association data in the KD Atlas would reveal molecular pathways through which gut-derived metabolites might influence CKD risk.

To investigate this hypothesis, we selected five gut microbiome-derived metabolites as input to the KD Atlas: hippurate (derived from dietary polyphenols metabolized by gut bacteria), phenylacetylglutamine (from phenylalanine metabolism) and three tryptophan-derived indole compounds (indoleacetate, indolelactate and indolepropionate). These metabolites were chosen because they represent key gut microbiome-derived metabolites linked to aromatic amino acid and polyphenol metabolism, and are known to be altered in CKD patients [89,90]. We constructed a metabolite-centric subnetwork by querying these five metabolites while expanding the network by 1-step using metabolite co-abundance (partial correlation data) to include additional metabolites. For network construction, we applied genome-wide significance to retain only robust mQTL connections. Genetic co-regulation edges were filtered for the ‘Kidney Cortex’ to focus on kidney-relevant associations.

The resulting network contains 129 protein-coding genes, 12 metabolites and 23 kidney disease traits (**Figure 5A**). Notably, the network expansion from 5 input metabolites to 12 total metabolites successfully captured two of the most important uremic toxins: indoxyl sulfate (3-indoxyl sulfate) and p-cresol sulfate, which were not in the original input set. This demonstrates the power of the network approach to identify biologically related metabolites. Additional metabolites in the expanded network also included tryptophan (the precursor for indole compounds), catechol sulfate, cyclo(pro-val), and isobutyrylcarnitine. Remarkably, 67 of the 129 genes in the network (52%) harbor genetic variants significantly associated with kidney disease traits, highlighting that the metabolite-gene connections reflect genuine biological pathways linking gut microbial metabolism to kidney disease. The 23 connected kidney disease traits span the full spectrum of kidney function assessment, including eGFRcrea, eGFRcys, diabetes-stratified analyses, BUN, UACR, CKD diagnosis, and measures of kidney function decline.

**Figure 5.**
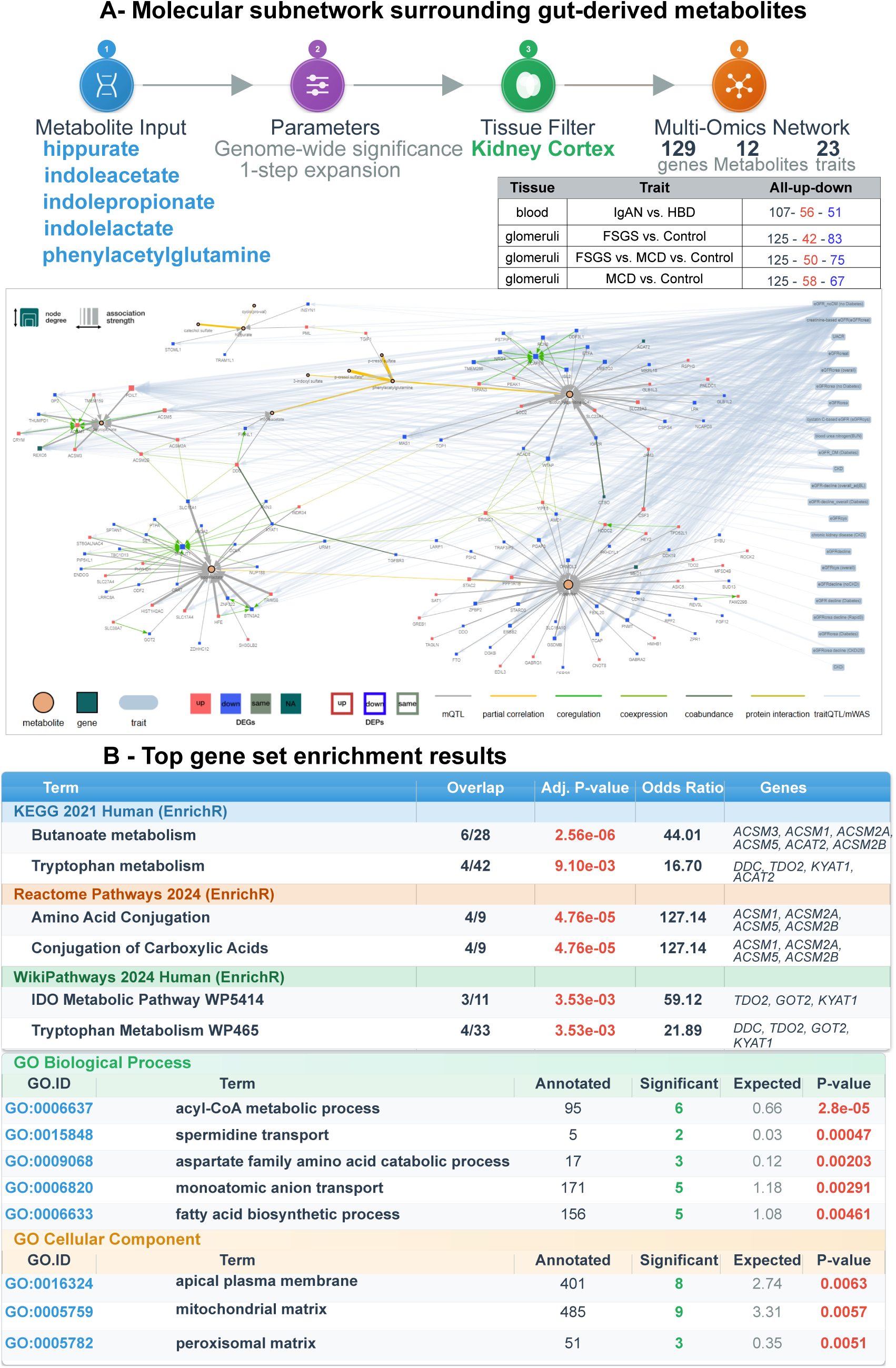
A genetic network links gut microbiome-derived metabolites to glomerular disease. **A.** Multi-omic subnetwork surrounding five gut-derived metabolites (hippurate, phenylacetylglutamine, indoleacetate, indolelactate, indolepropionate) from the KD Atlas. Node colors represent differential gene expression in glomeruli for the FSGS vs. MCD vs. Control comparison (red: up-regulated, blue: down-regulated). **B.** Enrichment analysis across multiple databases (KEGG 2021, Reactome 2024, WikiPathways 2024 via EnrichR; GO Biological Process and Cellular Component via topGO) reveals the network’s role in butanoate metabolism, amino acid conjugation, tryptophan metabolism, and organic cation transport. **Abbreviations**: FSGS, Focal Segmental Glomerulosclerosis; MCD, Minimal Change Disease.

Analysis of the 67 trait-associated genes revealed distinct functional categories that may provide mechanistic insights into how gut-derived metabolites influence kidney disease (**Figure 7A**). The most prominent categories include:

- Organic cation and anion transporters (*SLC22A1*, *SLC22A3*, *SLC17A1*, *SLC17A4*): These solute carrier family members are responsible for the tubular secretion of organic compounds, including uremic toxins. *SLC22A1* (OCT1) and *SLC22A3* (OCT3) encode organic cation transporters, while *SLC17A1* and *SLC17A4* encode organic anion transporters [93, 94]. Genetic variation in these transporters would be expected to alter the efficiency of uremic toxin elimination, thereby modulating systemic exposure to these compounds and influencing CKD progression risk.
- Acyl-CoA synthetases (*ACSM1*, *ACSM2A*, *ACSM2B*, *ACSM3*, *ACSM5*): This family of enzymes catalyzes the activation of medium-chain fatty acids and other organic acids by conjugating them with coenzyme A, enabling their participation in metabolic pathways [95]. The presence of ACSM family members in the network suggests that amino acid and organic acid conjugation pathways play a role in processing gut-derived metabolites. Genetic variants affecting these enzymes could alter the capacity for metabolite activation and downstream metabolism, potentially modulating systemic exposure to gut-derived compounds in CKD.
- Tryptophan and aromatic amino acid metabolism enzymes (*TDO2*, *KYAT1*, *DDC*, *GOT2*, *CRYM*): *TDO2* (tryptophan 2,3-dioxygenase) catalyzes the rate-limiting step in tryptophan catabolism via the kynurenine pathway [96]. *KYAT1* (kynurenine aminotransferase) acts downstream in this pathway. *DDC* (DOPA decarboxylase) is involved in aromatic amino acid metabolism. The enrichment of tryptophan metabolic enzymes directly connects to the indole metabolites in the network, as tryptophan is the precursor for indole compound synthesis by gut bacteria.
- Signaling and growth factors (*ERBB2*, *NRG4*, *FTO*, *CSF3*, *MED1*, *CDK12*), and mitochondrial and metabolic genes (*SOD2*, *ETFA*, *ACAD8*, *ACAT2*, *MRPL18*), point to specific pathways of cellular injury – oxidative stress, altered energy metabolism, and disrupted glomerular signaling – that may be activated by gut-derived metabolites.

To validate the functional coherence of the network, we performed enrichment analyses across KEGG, Reactome, WikiPathways and Gene Ontology databases within the KD Atlas. The results were highly consistent, supporting a coordinated genetic architecture linking gut microbial metabolism to kidney disease:

**KEGG pathway** analysis identified butanoate metabolism as the most significantly enriched pathway (see table, **Figure 5B**). Butyrate is a short-chain fatty acid produced by gut bacteria through fermentation of dietary fiber and has important roles in maintaining gut barrier function and modulating inflammation [97,98]. The strong enrichment of this pathway connects the network to gut microbial metabolism. Additional significantly enriched KEGG pathways included tryptophan metabolism, tyrosine metabolism and phenylalanine metabolism, directly validating the connection to aromatic amino acid catabolism that produces the indole and phenolic metabolites in the network.

**Reactome pathway** analysis provided even stronger validation, with amino acid conjugation emerging as the top enriched pathway. An identical result was obtained for conjugation of carboxylic acids, which represents the same biochemical process. These pathways describe the Phase II detoxification mechanism by which organic acids, including gut-derived metabolites, are conjugated with amino acids (primarily glycine) to enhance their water solubility and facilitate excretion [99]. As shown in the left table in **Figure 5B**, genes involved in metabolite detoxification are highly concentrated in this network. Additional Reactome pathways included metabolism of amino acids and derivatives, abacavir transmembrane transport and organic cation transport, highlighting the role of organic cation transporters in handling these metabolites.

**WikiPathways analysis** identified the IDO metabolic pathway and tryptophan metabolism as top enriched pathways (see table**, in Figure 5B**). IDO (indoleamine 2,3-dioxygenase) and TDO2 are the two enzymes that catalyze the initial step of tryptophan catabolism, producing kynurenine [100, 101]. This enrichment validates that the network captures tryptophan metabolic processes that generate indole compounds from unabsorbed dietary tryptophan.

**Gene Ontology enrichment analysis** revealed highly significant functional categories that align with the biochemical processes identified in pathway analyses (right table in **Figure 5B**). The most significantly enriched **GO biological processes** were acyl-CoA metabolic process, spermidine transport and metabolism, aspartate family amino acid catabolism, monoatomic anion transport and fatty acid biosynthetic process. **GO cellular component analysis** identified mitochondrial matrix, apical plasma membrane (which is the location of brush border transporters) and peroxisomal matrix. The enrichment of apical plasma membrane is particularly relevant, as this is where the *SLC22* family organic cation/anion transporters localize to mediate tubular secretion of uremic toxins [93].

Beyond the genetic architecture, we examined whether genes in the network show dysregulation at the transcript level in kidney disease. Differential gene expression analysis across multiple disease comparisons revealed widespread transcriptional changes in glomerular diseases (see table, upper right of **Figure 5**). In glomerular tissue comparisons, the network genes showed extensive dysregulation: focal segmental glomerulosclerosis (FSGS) versus controls displayed 125 differentially expressed genes (DEGs), with 42 up-regulated and 83 down-regulated; FSGS versus minimal change disease (MCD) versus controls showed 125 DEGs (50 up, 75 down); MCD versus controls had 125 DEGs (58 up, 67 down). In blood samples from IgA nephropathy (IgAN) patients, 107 DEGs were observed (56 up, 51 down). The consistent dysregulation of network genes across multiple distinct glomerular diseases – FSGS, MCD, and IgAN – suggests that these genes participate in common pathological processes affecting the glomerulus. Although previous work has established that indoxyl sulfate can induce free radical production in glomerular mesangial cells [91], the broader pathophysiological connection between gut-derived metabolite pathways and glomerular injury remains underexplored, with the literature predominantly focusing on tubular toxicity and tubular epithelial cell dysfunction [89, 91]. Our finding that a unified genetic network linking gut microbial metabolites shows widespread and coordinated dysregulation across multiple glomerular diseases (FSGS, MCD, IgAN) therefore reveals a potential novel, genetically modulated pathway. This provides a mechanistic architecture for how gut-derived metabolites may contribute to glomerular injury and disease progression. The presence of *ERBB2* and *NRG4* in this network offers a specific mechanistic hypothesis, as *ERBB* signaling pathways are critical for podocyte health and glomerular filtration barrier integrity [102,103], suggesting a potential link beyond previously reported mesangial cell toxicity. Systemic exposure to uremic toxins [91,104] modulated by genetic variation in metabolite-processing enzymes (ACSM family[105], *TDO2*, *KYAT1* [100]) and transporters (SLC22 family) [93], may influence glomerular disease risk through effects on podocyte signaling pathways [103,106].

Integration of these findings leads to a mechanistic model for how genetic variation in metabolite-processing pathways influences CKD risk: (1) Gut microbiota metabolize dietary tryptophan and other aromatic amino acids, producing indole derivatives, p-cresol derivatives, and other compounds [89,107–108]. (2) Genetic variation in tryptophan catabolism enzymes (*TDO2*, *KYAT1*, *DDC*) modulates the systemic availability of tryptophan that reaches the colon, thereby influencing substrate availability for microbial metabolism [109,110]. (3) Absorbed gut-derived metabolites undergo Phase II detoxification via amino acid conjugation (ACSM family enzymes), and genetic variants in these enzymes can alter detoxification capacity. For example, a stop-gain variant in *ACSM2A* impairs the conjugation of the gut-derived tryptophan metabolite indolepropionic acid, leading to its substantial systemic accumulation [111]. 4) Conjugated metabolites are secreted into urine by organic cation and anion transporters (*SLC22A1*, *SLC22A3*, *SLC17A1*, *SLC17A4*) in the proximal tubule, with genetic variants affecting secretion efficiency and thus determining systemic exposure to uremic toxins [93,112]. (5) Elevated systemic exposure to uremic toxins affects multiple kidney compartments, including tubular epithelial cells (direct toxicity, oxidative stress reflected by *SOD2*) [113] and glomerular podocytes (potentially through *ERBB2*/*NRG4* signaling), contributing to both tubular dysfunction and glomerular disease [103]. (6) Over time, these effects contribute to progressive CKD, with the extent of kidney function decline determined partly by individual genetic variation in the metabolite-processing pathway.

In conclusion, this analysis demonstrates how the KD Atlas enables hypothesis-generating research by revealing unexpected molecular connections. Starting from five gut microbiome-derived metabolites, we constructed a network that: (1) successfully expanded to include major uremic toxins (indoxyl sulfate, p-cresol sulfate), (2) identified 67 genes (52% of the network) with genetic associations to kidney function, (3) revealed coherent enrichment of amino acid conjugation, tryptophan metabolism and organic cation transport pathways and (4) showed unexpected connections to glomerular disease through widespread dysregulation in FSGS, MCD and IgAN. The presence of *ERBB2* and *NRG4* in this network suggests that genetic variation in metabolite-processing enzymes and transporters may modulate individual susceptibility to uremic toxicity through effects on both tubular and glomerular compartments. This showcase illustrates the power of multi-omics network integration to generate novel mechanistic hypotheses that bridge gut microbiome metabolism, genetic architecture, and kidney disease pathology.

## 4. Discussion

We present the KD Atlas, a comprehensive network-based resource that provides a global, multi-omics view of kidney disease. By adapting and extending the technical framework of the AD Atlas [31], we demonstrate the generalizability of this QTL-based integration approach for complex diseases beyond neuroscience. The KD Atlas integrates data from over 25 studies into a unified knowledge resource, enabling researchers to dynamically generate and explore context-specific molecular subnetworks through an intuitive web interface (https://metabolomics.helmholtz-munich.de/kdatlas). For the construction of the KD Atlas leveraging the existing framework, we followed a two-part strategy: retaining and extending the disease-agnostic infrastructure, including knowledge databases and population-based data, while comprehensively replacing AD-specific associations with kidney disease-specific datasets.

The resulting resource provides access to more than 20,000 protein-coding genes, 1,300 metabolites, nearly 2,000 proteins and 40 kidney disease traits connected by over 1.2 million relationships. Through the interactive interface, researchers can dynamically construct molecular subnetworks focused on their own research questions by using three primary entry points – trait-centric, gene-centric and metabolite-centric – and tailor them with tissue-specific filters to focus on kidney-relevant relationships. Users can overlay differential expression data and perform functional enrichment analyses on these subnetworks without requiring specialized bioinformatics expertise. The KD Atlas thus maintains the powerful, user-friendly features of the AD Atlas while offering dedicated analytical capabilities tailored to renal physiology and pathology.

In two showcases, we demonstrated that the KD Atlas is a powerful tool for highlighting and contextualizing genetic associations, using examples for which this context and the relevance for kidney health is well established. The subnetworks surrounding *UMOD* and *CUBN* did not merely list associated genes; they reconstructed functionally coherent pathways and protein complexes. The *UMOD* network successfully captured the thick ascending limb ion transport machinery, connecting uromodulin to key transporters such as *SLC12A1* (NKCC2) and *CASR*, with enrichment analyses highlighting potassium ion homeostasis and chloride transport [73, 74]. Similarly, the *CUBN* network automatically identified the essential *CUBN*-*LRP2*-*AMN* receptor complex and highlighted enrichment of cobalamin transport genes in the subnetwork, providing a systems-level explanation for its role in vitamin B12 reabsorption and proteinuria [85, 86]. This ability to embed genetic findings within their functional molecular neighborhoods addresses a critical need in post-GWAS research.

Beyond extracting known biology, the KD Atlas excels at hypothesis generation. The analysis of gut microbiome-derived metabolites revealed a previously less appreciated genetic architecture. Starting from five metabolites, network expansion captured major uremic toxins and connected them to 129 genes, 67 of which (52%) are associated with kidney disease traits. The most unexpected finding was the widespread dysregulation of these genes across diverse glomerular diseases (FSGS, MCD, IgAN), suggesting a shared pathophysiological mechanism [55, 57]. The presence of *ERBB2* and *NRG4,* which are critical for podocyte health [102, 103], in this network of metabolite-processing enzymes and transporters suggests a novel hypothesis: that systemic exposure to gut-derived metabolites, modulated by genetic variation, may influence glomerular disease risk through effects on podocyte signaling, moving beyond the established paradigm of direct tubular toxicity [89, 104].

The kidney research community is served by several valuable omics resources, which have been comprehensively reviewed elsewhere [25]. These can be broadly categorized by their primary data focus. For transcriptomics, Nephroseq integrates gene expression in renal disease with clinical data across 26 datasets, while the Renal Gene Expression Database provides an easy-to-use interface covering 88 research papers on gene expression in renal disease. Specialized proteomics databases offer depth in protein cataloguing, including the Human Kidney and Urine Proteome Project (HKUPP) for protein expression in normal urine and kidney tissues, and the Urinary Protein Biomarker Database (UPBD), which contains over 400 reports on candidate protein biomarkers. For peptidomics, the Urinary Peptidomics and Peak-maps Database focuses on urinary peptides modified in disease. Multi-omics resources such as the Kidney and Urinary Pathway Knowledge Base (KUPKB) and the Chronic Kidney Disease Database (CKDdb) provide curated collections of publicly available datasets from various molecular levels, with KUPKB comprising over 220 experiments and CKDdb housing 366 datasets searchable by study, sample, tissue, disease, and molecule type. It should be noted that as of 2025, several databases mentioned in the original 2016 review (including UPDB, KUPKB, and CKDdb) appear to be no longer accessible, highlighting ongoing challenges in long-term database maintenance within the bioinformatics field.

Two further resources of particular importance to the kidney disease community deserve explicit mentioning: First, the Kidney Precision Medicine Project (KPMP) provides a large-scale, openly accessible atlas of human kidney cell types in health and disease, integrating single-cell and single-nucleus RNA sequencing, spatial transcriptomics, epigenomics and proteomics from carefully phenotyped kidney biopsy samples of patients with AKI and CKD, with all data freely available at kpmp.org [114]; notably, bulk RNA-seq data from KPMP were used in the present work to derive kidney tissue-specific gene co-expression networks integrated into the KD Atlas. Second, the Susztak Laboratory Kidney Biobank (susztaklab.com) hosts a comprehensive collection of human kidney multi-omics datasets (encompassing bulk and cell-type-resolved expression QTL atlases across tubular and glomerular compartments, methylation QTL, protein QTL, single-nucleus chromatin accessibility, single-cell transcriptomics and spatial transcriptomics) that have underpinned a series of landmark studies connecting genetic variants to kidney disease mechanisms at cell-type resolution [19,20,21,24].

The KD Atlas introduces a distinct paradigm compared to these existing resources. While the aforementioned databases are invaluable as repositories and for focused queries, the KD Atlas is built around a dynamic, network-based integration model. Its core innovation lies in a unified graph database that not only aggregates data but also enables on-demand construction of context-specific molecular subnetworks. This supports a systems-level perspective, allowing researchers to move from a list of associated entities to an interactive network view that connects genetic variants, transcript co-expression, protein co-abundance, metabolic correlations, and protein–protein interaction data within a single framework. Unlike the Kidney Disease Genetic Scorecard [24], which integrates 32 genomic annotation types to systematically prioritize causal variants and their target genes within the genetic-regulatory axis, the KD Atlas is designed for open-ended, cross-layer exploratory analysis: users can enter any gene, metabolite or kidney disease trait as a starting point and immediately interrogate its molecular context across all integrated omics layers, without requiring bioinformatics expertise.

The current version of the KD Atlas has several limitations. First, the molecular framework is largely data-driven. While it incorporates experimentally supported protein-protein interactions from databases such as STRING [31], HIPPIE [32] and IID [33], integration of additional expert-curated pathways and drug-target knowledge would further enhance biological interpretation and contextual relevance. Second, our genetic data predominantly derive from European-ancestry populations, which limits generalizability. As genetic associations and regulatory relationships vary across populations, future updates should prioritize the inclusion of trans-ethnic datasets to capture ancestry-specific mechanisms. Third, differential expression data coverage across kidney diseases is currently incomplete. Studies focusing on specific kidney compartments (e.g., cortex, medulla), as well as transcriptomic profiles from focal segmental glomerulosclerosis, minimal change disease and IgA nephropathy, have been integrated, but additional contexts, such as membranous nephropathy, lupus nephritis and specific diabetic kidney disease subtypes, will further improve comprehensiveness. Future updates will incorporate these and other relevant datasets as they become available. Fourth, association networks in the KD Atlas are constructed from datasets with diverse designs, cohort sizes, and technical platforms. Selection of significance thresholds is necessary to avoid overly dense networks, and trade-offs exist between false positive and false negative associations. While the underlying database contains fine-grained associations, these are currently summarized using study-specific thresholds. Systematic meta-analysis or cross-study weighting has not yet been implemented. As more high-quality datasets become available, future updates will adopt standardized processing, harmonization and context-dependent edge weighting to reduce bias and enhance reliability across studies. Integration of human kidney multi-omics datasets hosted at the Susztak Laboratory Kidney Biobank (susztaklab.com), including expression QTL atlases across tubular and glomerular compartments, protein QTL and spatial transcriptomics data [19,20,21,24], represents a priority direction for future development, with the potential to substantially enhance the genetic and molecular resolution of the KD Atlas. Future development will introduce graph-based module detection, allowing users to automatically identify densely connected functional communities within their subnetworks. This will facilitate the discovery of novel disease-relevant biological processes by enabling the separate functional characterization of these distinct modules.

## 5. Conclusion

The KD Atlas provides a comprehensive multi-omics resource for kidney disease interactively explorable at https://metabolomics.helmholtz-munich.de/kdatlas. By integrating data from over 25 studies into a unified network, it enables the construction of context-specific molecular subnetworks around user-selected genes, metabolites, and kidney disease traits. As shown in proof-of-concept analyses, generated subnetworks reflect established kidney disease biology. Additionally, the tailored, dynamic subnetwork extraction supports the generation of novel mechanistic hypotheses and may help in prioritization of therapeutic targets. Its network-based structure integrating multiple omics layers provides unique insights into coordinated molecular processes, establishing a foundational resource for future research into kidney disease mechanisms.

## Data availability

The KD Atlas is accessible via the user interface at https://metabolomics.helmholtz-munich.de/kdatlas. A comprehensive listing of the exact data sources integrated in the KD Atlas is given in the **Supplementary Material**.

## Acknowledgement

We thank the providers of kidney disease-related data used in our study, including data available from the CKDGen Consortium repository (https://ckdgen.imbi.uni-freiburg.de) and Nephroseq (https://nephroseq.org). Also, our results are in part based upon data generated by the Kidney Precision Medicine Project. Accessed December 18, 2023. https://kpmp.org. The Kidney Precision Medicine Project (KPMP) is supported by the National Institute of Diabetes and Digestive and Kidney Diseases (NIDDK) through the following grants: U01DK133081, U01DK133091, U01DK133092, U01DK133093, U01DK133095, U01DK133097, U01DK114866, U01DK114908, U01DK133090, U01DK133113, U01DK133766, U01DK133768, U01DK114907, U01DK114920, U01DK114923, U01DK114933, U24DK114886, UH3DK114926, UH3DK114861, UH3DK114915, and UH3DK114937. We gratefully acknowledge the essential contributions of our patient participants and the support of the American public through their tax dollars. This work was supported by the de.NBI Cloud within the German Network for Bioinformatics Infrastructure (de.NBI) funded by the German Federal Ministry of Education and Research (BMBF) (031A532B, 031A533A, 031A533B, 031A534A, 031A535A, 031A537A, 031A537B, 031A537C, 031A537D, 031A538A). MA and GK have been supported by several grants from the National Institute on Aging (NIA) within the funding scheme of the US National Institutes of Health (NIH): RF1AG057452, RF1AG058942, RF1AG059093, U01AG061359, U19AG063744, R01AG069901, R01AG081322. GK and YANN received support from the Deutsche Forschungsgemeinschaft (DFG), FOR 5795 (Projektnr. 536691227). This work was also supported by the German Federal Ministry of Education and Research (BMBF) within the framework of the e:Med research and funding concept (grant number: 01ZX1912D). The work of SH and AK was supported by the DFG Project ID431984000 SFB 1453.

## Author Contributions

Conceptualization: MA, GK; data analysis and resource development: YANN, MAU, OB, MA, GK; investigation: YANN, SH, AK; visualization: YANN, MAU; software: WRM; funding acquisition: MA, GK; supervision: JD, MA, GK; writing - original draft: YANN, GK; writing – review: all authors read and approved the final manuscript.

