## Supplementary Material for "The KD Atlas: A Multi-Omics Network Resource for Kidney Disease Research"

### 1. Framework Overview: Adaptation of the AD Atlas to Kidney Disease

#### 1.1 Relationship to the Alzheimer's Disease Atlas

The Kidney Disease Atlas (KD Atlas) builds upon the technical infrastructure and integration methodology established by the Alzheimer's Disease (AD) Atlas [1]. The AD Atlas framework was explicitly designed to be generalizable across complex diseases through its disease-agnostic core architecture: a quantitative trait locus (QTL)-based integration strategy paired with a composite network approach that enables systematic integration of multi-omics data without requiring matched samples across all modalities [1,2].

Our development strategy involved retaining the entire core infrastructure – including the data model, the two-phase integration pipeline (data collection/integration followed by abstraction) and the web interface framework—while systematically replacing all disease-specific association data (**Figure S1**). Specifically, we removed all AD GWAS, brain-specific differential expression analyses, and AD-related metabolome-wide association studies, replacing them with kidney disease equivalents. Additionally, we extended the resource by integrating protein-protein interaction networks from established knowledge databases and novel population-based datasets not present in the original AD Atlas. Core methodological details that remain unchanged from the AD Atlas are described previously [1]. This supplement provides complete documentation of all kidney-specific data collection, curation, and processing workflows that constitute the novel contributions of this work.

#### 1.2 Integration Strategy

The KD Atlas implements a step-wise multi-omics integration approach described previously [1]. This methodology facilitates the consolidation of heterogeneous datasets originating from diverse cohorts, studies and analytical platforms without requiring matched samples across all omics modalities. Networks are constructed by establishing pairwise relationships categorized as inter-omics, intra-omics and phenotype-specific links (Section 1.3), utilizing both knowledge-based databases and data-driven statistical associations. These relationships connect biological entities through overlapping quantitative trait loci (QTLs), ultimately forming a composite network architecture stored and queried within the Neo4j graph database system.

#### 1.3 Inferred Relationship Types

Three distinct categories of molecular relationships were integrated into the KD Atlas (**Figure S1**):

**Intra-omics relationships** connect entities within a single omics layer by leveraging the inherent correlation structures of molecular datasets. For the KD Atlas, we integrated kidney tissue gene co-expression networks establishing gene–gene links, as well as partial correlation-based networks representing protein co-abundance and metabolite co-occurrence patterns (protein–protein and metabolite–metabolite links, respectively).

**Inter-omics relationships** bridge distinct omics layers through two complementary strategies. First, knowledge-based mappings from Ensembl establish direct connections between genes, transcripts and proteins. Second, quantitative trait locus (QTL) data enable linkage across layers by identifying shared genetic determinants. The KD Atlas incorporates tissue-specific expression QTL (eQTL) data from GTEx version 8 covering 49 tissues, including kidney cortex. Additionally, protein QTLs (pQTLs) and metabolite QTLs (mQTLs) from population-based cohorts establish genetic connections to proteomic and metabolomic measurements.

**Phenotype-specific relationships** link molecular entities to kidney disease traits. These associations are derived from large-scale genome-wide association studies (GWAS) and metabolome-wide association studies (MWAS) investigating kidney disease phenotypes. This layer enables identification of genes, proteins and metabolites implicated in kidney disease pathophysiology (comprehensive list in **Table S2**).

#### 1.4 Data Replacement and Extension Strategy

The AD Atlas [1] was designed as a flexible framework that facilitates transfer to other disease contexts, enabling the systematic replacement of AD- and brain-specific components with kidney disease- and kidney tissue-specific equivalents. All datasets unrelated to AD – specifically those derived from knowledge databases and population-based studies – were retained without modification, as they are disease-agnostic and had already been preprocessed within the AD Atlas framework [1]. These include gene–transcript–protein mappings, SNP annotations, and population-level QTL and correlation data.

In contrast, AD-specific datasets, including GWAS, MWAS, differential expression data and co-expression networks, were replaced with kidney disease-specific datasets curated and processed. This replacement incorporated large-scale genome- and metabolome-wide association studies, as well as kidney omics and co-expression networks derived from kidney disease cohorts.

Beyond replacing disease-specific components, the KD Atlas integrates additional population-based datasets not present in the AD Atlas, including protein co-abundance networks from the UK Biobank and experimentally validated protein–protein interaction networks, broadening the range of functional and molecular relationships. **Table S4** summarizes the data categories in the KD Atlas: those retained from the AD Atlas, those replaced with kidney disease-specific equivalents, and newly added resources.

### 2. Data

The KD Atlas integrates diverse datasets derived from knowledge databases, population-based studies and disease-specific sources. Disease-agnostic datasets described previously [1] were retained without modification, while AD-specific datasets were replaced with kidney disease-specific equivalents. Additionally, newly available resources were incorporated to expand network coverage.

#### 2.1 Public Databases and Population-Based Studies

The following disease-agnostic resources from the AD Atlas [1] were retained: gene-transcript-protein mappings (Ensembl [3]), SNP annotations (SNiPA [4]), tissue-specific eQTL data (GTEx v8 [5]), five mGWAS studies [6-10], and two partial correlation network studies [11, 12] (**Table S4**). Detailed methodology for these datasets is described previously [1].

Beyond these retained resources, we integrated one additional partial correlation network study and three protein-protein interaction databases.

##### 2.1.1 Partial correlation networks

**Suhre et al., 2024 [13]** – Genetic associations with ratios between protein levels using UK Biobank proteomics data (>54,000 samples, 1,463 proteins measured with the Olink platform). The study identified 4,248 replicated associations with 2,821 protein ratios (rQTLs), representing a 24.7% increase in discovered genetic signals compared to analyses of absolute protein levels alone. A total of 11,936 Bonferroni-significant Gaussian Graphical Model (GGM) edges used for rQTL testing were downloaded and integrated into the KD Atlas.

##### 2.1.2 Protein-Protein Interaction Networks

To construct a high-confidence human protein-protein interaction (PPI) network, we integrated experimentally validated physical interactions from three major sources:

STRING (v11.0) [14], HIPPIE (v2.0) [15] and IID (v2021.05) [16]. Gene identifiers were harmonized across all sources by mapping to Ensembl IDs (GRCh38 p13), ensuring consistency and enabling reliable integration. Interactions were filtered for high confidence using database-specific thresholds (HIPPIE ≥0.73; STRING ≥0.7), resulting in 170,661 PPIs connecting 14,991 unique proteins. To further assess interaction reliability, PPIs were classified according to their recurrence across sources: high-confidence interactions (n = 54,147) were supported by all three databases, medium-confidence interactions (n = 85,269) were present in two, and low-confidence interactions (n = 31,245) were reported in a single database. This integrative approach generated a robust, confidence-stratified human PPI network that was incorporated into the KD atlas, enabling users to explore protein interactions at varying levels of experimental support.

#### 2.2 Kidney Disease-Specific Data Collection

In the following, we list the kidney disease-specific datasets integrated in the current version of the KD Atlas, along with their sources.

##### 2.2.1 Genome-Wide Association Studies for Kidney Function and Disease

GWAS summary statistics for kidney function and disease phenotypes were obtained primarily from the Chronic Kidney Disease Genetics (CKDGen) Consortium [17]. Only summary statistics from individuals of European ancestry were included. Most data were downloaded from the CKDGen repository, with the exception of two studies (Winkler et al., 2022 [18]; Gorski et al., 2022 [19]), which were obtained from alternative sources.

**Winkler et al., 2022 [18]** – Diabetes-stratified GWAS meta-analysis of estimated glomerular filtration rate (eGFR) based on serum creatinine. Genetic determinants were analyzed separately in individuals with diabetes mellitus (n = 178,691) and without diabetes (n = 1,296,113). The study identified seven loci with significant DM/noDM differences, four additional loci with suggestive differences, and 28 novel loci when allowing for potential differences. Analyses used standard additive models adjusted for age, sex, and ancestry principal components.

**Gorski et al., 2022 [19]** – Meta-analysis of 62 longitudinal studies (n = 343,339) examining genetic loci associated with annual eGFR decline, a key predictor of progression to kidney failure. Twelve genome-wide significant independent variants were identified (11 novel for the decline phenotype), with several showing variant-by-age interactions and two- to four-fold greater effects in high-risk subgroups. Analyses included unadjusted models and models adjusted for baseline eGFR.

**Stanzick et al., 2021 [20]** – Large-scale meta-analysis combining CKDGen Consortium and UK Biobank data (n = 1,201,909 for creatinine-based eGFR) to identify genetic loci associated with kidney function. The study reported 424 loci for eGFRcrea (201 novel), explaining 9.8% of eGFRcrea variance via 634 independent signals. Fine-mapping resolved 99% credible sets to single variants for 44 signals and ≤5 variants for 138 signals, improving causal variant prioritization. Cystatin C-based eGFR (n = 460,826) and blood urea nitrogen (n = 852,678) provided complementary phenotypes supporting 348 loci.

**Gorski et al., 2021 [21]** – Meta-analysis of 42 studies examining rapid kidney function decline, a phenotype associated with severe clinical endpoints. Two complementary definitions were used: Rapid3 (eGFR decline ≥3 mL/min/1.73 $m^2$/year; 34,874 cases vs 107,090 controls) and CKDi25 (≥25% eGFR decline plus incident eGFR <60 mL/min/1.73 $m^2$; 19,901 cases vs 175,244 controls). Seven independent variants across six loci were identified, all novel for rapid decline phenotypes. The OR2S2 locus was novel for any eGFR trait. Individuals with high genetic risk (8–14 adverse alleles) exhibited a 1.20-fold increased risk of acute kidney injury (95% CI 1.08–1.33).

**Tin et al., 2019 [22]** – Trans-ancestry GWAS of serum urate levels (n = 457,690) identifying 183 loci (147 novel) influencing urate metabolism and its connection to cardiometabolic traits and gout. Enrichment analyses highlighted the kidney and liver as key target organs. Experimental validation demonstrated HNF4A transactivation of ABCG2, a major urate transporter in kidney cells, and identified the HNF4A p.Thr139Ile variant as functional.

**Teumer et al., 2019 [23]** – Trans-ethnic meta-analysis of urinary albumin-to-creatinine ratio (UACR) and microalbuminuria (n = 564,257). Identified 68 UACR-associated loci. Fine-mapping and trans-omics analyses integrating gene expression across 47 tissues and plasma protein levels implicated genes acting through differential expression in kidney, including TGFB1, MUC1, PRKCI, and OAF. Functional validation in Drosophila nephrocytes demonstrated that knockdown of OAF and PRKCI orthologs reduces albumin endocytosis, and silencing PRKCI additionally impairs slit diaphragm formation.

**Wuttke et al., 2019 [24]** – Trans-ancestry meta-analysis of eGFR and related kidney traits (n = 1,046,070). Identified 264 loci associated with eGFR (166 novel), with 147 likely relevant for kidney function based on blood urea nitrogen associations (n = 416,178). Pathway enrichment analyses supported the kidney as the main target organ. Colocalization with gene expression across 46 tissues identified 17 genes differentially expressed in kidney compartments. Fine-mapping highlighted missense driver variants in 11 genes and kidney-specific regulatory variants.

**Gorski et al., 2017 [25]** – 1000 Genomes-based meta-analysis of kidney function (n = 110,517, European ancestry), identifying 10 novel loci for eGFR previously missed by HapMap-based GWAS. Six loci were tagged by common SNPs unique to the 1000

Genomes reference panel (HOXD8, ARL15, PIK3R1, EYA4, ASTN2, EPB41L3). Pathway analysis revealed 39 significant genes (FDR < 0.05) and 127 enriched gene sets (FDR < 0.05), including pathways related to kidney development, carbohydrate metabolism, and glucose metabolism.

**Pattaro et al., 2016 [26]** – Meta-analysis of eGFR and CKD (n = 133,413 discovery; n = 42,166 replication), identifying 24 new and confirming 29 previously reported loci (53 total). Nineteen loci were associated with eGFR in individuals with diabetes. Bioinformatic analyses revealed enrichment for kidney tissue expression and pathways related to kidney development, transmembrane transport, kidney structure, and glucose metabolism regulation. Chromatin state mapping and DNase I hypersensitivity showed preferential localization of associated variants to regulatory regions in kidney but not extra-renal tissues. Integrated trait: eGFRcrea.

**Teumer et al., 2015 [27]** – Diabetes-stratified GWAS meta-analysis of albuminuria in individuals with diabetes (n = 5,825) and without diabetes (n = 46,061). Confirmed associations at CUBN (encoding cubilin) with UACR in the overall sample ($P = 2.4 \times 10^{-10}$). Detected gene-by-diabetes interactions at HS6ST1 and RAB38/CTSC, with genetic effects observed only in individuals with diabetes (21% UACR change per minor allele for HS6ST1, $P = 6.3 \times 10^{-7}$; 13% for RAB38/CTSC, $P = 5.8 \times 10^{-7}$). Functional studies in streptozotocin-induced diabetic Rab38 knockout rats showed higher urinary albumin and reduced megalin/cubilin at the proximal tubule cell surface compared with controls.

**Gorski et al., 2015 [28]** – GWAS meta-analysis of kidney function decline in individuals of European descent (n = 63,558). SNPs at MEOX2, GALNT11, IL1RAP, NPPA, HPCAL1, CDH23, and the known UMOD locus showed strongest associations. UMOD reached genome-wide significance for annual eGFR change and was replicated. GALNT11 (associated with Rapid Decline) and CDH23 (associated with eGFR change in CKD) showed suggestive replication. Morpholino knockdowns of galnt11 and cdh23 in zebrafish resulted in severe edema after gentamicin exposure. Integrated traits: eGFR-decline, Rapid Decline, CKD-specific eGFR change.

##### 2.2.2 Metabolome-Wide Association Studies (MWAS)

The KD Atlas integrates metabolomic data from four large MWAS, comprising urine and serum measurements from cross-sectional and longitudinal studies. These studies examined associations between metabolites and kidney-related outcomes, including kidney function, CKD, kidney failure, acute kidney injury, and mortality. The datasets include non-targeted urine metabolomics from the German Chronic Kidney Disease (GCKD) study, non-targeted serum metabolomics from KORA F4 and TwinsUK, and targeted serum metabolomics from KORA S4/F4. Only metabolites that were statistically significant in the original studies were integrated into the Atlas.

**Steinbrenner et al., 2021 [29]** - GCKD study employing discovery-replication design (2:1 ratio) with n=5,087 CKD patients. Non-targeted urine metabolomics quantified 1,487 metabolites. Multivariable Cox regression (adjusted for age, sex, diabetes, eGFR, UACR, proteinuria) identified 55 metabolites significantly associated with outcomes after median 4-year follow-up. C-glycosyltryptophan showed consistent associations with all three endpoints (HR 1.43 for kidney failure, 1.40 for combined kidney failure and acute kidney injury, 1.47 for death). Metabolites in phosphatidylcholine pathway showed significant enrichment. Outcomes: kidney failure (241 events), KF+AKI (382 events), mortality (362 events).

**Peggy et al., 2016 [30]** - KORA F4 discovery (n≤1,735) with TwinsUK replication (n=1,164). Gas and liquid chromatography coupled to mass spectrometry quantified 493 serum metabolites. After multiple testing correction, 54 metabolites significantly associated with eGFRcr, with six showing strong correlation (r≥0.50) with established kidney function markers: C-mannosyltryptophan, pseudouridine, N-acetylalanine, erythronate, myo-inositol, N-acetylcarnosine. Higher C-mannosyltryptophan, pseudouridine, and O-sulfo-L-tyrosine associated with incident CKD. C-mannosyltryptophan and pseudouridine showed little sex dependence (unlike creatinine) and correlation with measured GFR of 0.78 in AASK study validation (n=200).

**Goek et al., 2013 [31]** - KORA S4/F4 longitudinal study (n=1,104, 7-year follow-up). Flow injection and liquid chromatography tandem mass spectrometry measured 140 metabolites and 19,460 metabolite ratios. After multiple testing correction, spermidine ($P=5.8\times10^{-7}$), phosphatidylcholine diacyl C42:5-to-phosphatidylcholine acyl-alkyl C36:0 ratio ($P=1.5\times10^{-6}$), and kynurenine-to-tryptophan ratio ($P=1.9\times10^{-6}$) significantly associated with annual eGFR change. Kynurenine-to-tryptophan ratio also associated with incident CKD (OR 1.36 per SD, 95% CI 1.11-1.66, $P=2.7\times10^{-3}$).

**Goek et al., 2012 [32]** - KORA F4 discovery (n=3,011) with TwinsUK validation (n=984). Targeted mass spectrometry quantified 151 metabolites and 22,650 metabolite ratios. Meta-analysis identified 22 replicated metabolites and 516 metabolite ratios associated with eGFR (pooled P ranging from $7.1\times10^{-7}$ to $1.8\times10^{-69}$ for single metabolites). Acylcarnitines such as glutarylcarnitine inversely associated with eGFR (-3.73 mL/min/1.73m² per SD, pooled $P=1.8\times10^{-69}$). Serine-to-glutarylcarnitine ratio showed strongest association ($P=3.6\times10^{-81}$). Almost all replicated phenotypes associated with decreased eGFR (<60 mL/min/1.73m²; n=172 cases) with ORs ranging 0.29-2.06 per SD.

##### 2.2.3 Genetic Associations with Metabolic Traits (mGWAS)

**Schlosser et al., 2023 [33]** – Genetic studies of paired plasma and urine metabolomes investigating enzymatic and transport processes at the interface of body compartments. A genome-wide association study of 1,916 metabolites measured in paired plasma and urine samples from the German Chronic Kidney Disease (GCKD) study identified 1,299 significant genetic associations, with 40% of implicated metabolites detectable only in urine analyses, highlighting kidney-specific metabolic processes not captured by plasma-only studies. Summary statistics were downloaded and integrated into the KD Atlas.

##### 2.2.4 Differential Gene Expression Analysis in Kidney Disease Cohorts

Differential gene expression data were obtained primarily from the Nephroseq database (https://nephroseq.org/), a publicly available repository of kidney-related expression datasets. Selection criteria included: (1) human samples, (2) reported European ancestry demographics, and (3) availability of differential analysis results. One additional study (Haug et al., 2024 [34]) was identified through literature search.

**Haug et al., 2024 [34]** - Multi-omic analysis of human kidney cortex versus medulla using tumor-adjacent normal tissue from five male nephrectomy patients. Integrated RNA-seq, ATAC-seq, Hi-C, and spatial transcriptomics to characterize medulla-specific gene expression patterns, chromatin accessibility signatures, and three-dimensional chromatin organization. Used carefully annotated specimens to reassign incorrectly labeled samples in GTEx Project and extract meaningful medullary gene expression signatures. Validated findings through spatial transcriptomics and immunohistochemistry. Datasets provide resource for functional annotation of GWAS variants and are accessible through epigenome browser portal. Comparison: Cortex vs. Medulla. Tissue: Normal kidney. Platform: RNA-seq with epigenomic validation.

**Cox et al., 2015 [35]** - Genome-wide expression analysis of peripheral blood monocytes from IgA nephropathy patients (n=39) versus healthy donors (n=37). Microarray analysis (with RT-PCR and flow cytometry validation) identified altered apoptosis signaling, mitochondrial dysfunction, and death receptor pathways. Protein levels of NDUFS3 and TNFRSF1A upregulated, confirming altered mitochondrial and death receptor homeostasis. Basal intracellular TNF protein levels lower in IgAN patients. Non-classical monocyte subset ($CD14^{+}CD16^{+}$) significantly expanded in all IgAN patients despite unchanged total monocyte count. Comparison: IgAN vs. healthy blood donors (HBD). Tissue: Peripheral blood monocytes. Platform: Microarray.

**Hodgin et al., 2010 [36]** - Molecular profiling of focal segmental glomerulosclerosis using laser-captured glomeruli from formalin-fixed paraffin-embedded renal biopsies. Affymetrix array analysis of mRNA isolated from archived biopsies (recently considered unsuitable for microarray but successfully utilized here). Unsupervised hierarchical clustering revealed two distinct clusters delineating FSGS and collapsing FSGS from normal controls and minimal change disease. Class comparison identified 316 differentially regulated genes (134 up, 182 down) in FSGS+collapsing FSGS versus normal+MCD. Genes included slit diaphragm junctional complex components and previously described dysregulated podocyte phenotype markers. Gene Ontology analysis revealed overrepresented processes: development, differentiation, morphogenesis, cell motility/migration, cytoskeleton organization, signal transduction. Transcription factors associated with developmental processes heavily overrepresented. Comparisons: FSGS vs. control, collapsing FSGS vs. control, FSGS vs. MCD, MCD vs. control. Tissue: Glomeruli. Platform: Affymetrix Human X3P array.

**Rodwell et al., 2004 [37]** - Transcriptional profiling of aging in human kidney cortex and medulla. Identified 985 genes changing expression with age in both regions. Age-regulated genes showing similar aging profile in cortex and medulla suggested common underlying aging mechanism. Genes with increasing transcript abundance included those specifically expressed in immune cells, suggesting increased immune surveillance or inflammation with age. Expression profiles of age-regulated genes marked not only chronological age but also relative health and physiology of kidney in older individuals. Comparisons: Age-related changes in cortex and medulla; proteinuria abnormal vs. normal (separate for cortex and medulla). Tissue: Kidney cortex and medulla. Platform: Microarray.

##### 2.2.5 Differential Protein Abundance Analysis in Kidney Disease Cohorts

**Glorieux et al., 2015 [38]** - High-resolution plasma proteome analysis comparing CKD stages. Discovery phase: stage 2-3 CKD patients (n=14) versus stage 5 patients on hemodialysis (HD) (n=15). Validation cohort: 40 patients across CKD stages with/without HD. LC-MS/MS analysis detected 2,054 proteins; 333 showed differential abundance (127 decreased, 206 increased in HD patients, p-values not explicitly stated but inferred from statistical methods). Molecular pathway analysis confirmed known CKD complications: decreased hemostasis, increased inflammation, complement activation, vascular damage. Identified plasma increase during CKD progression of lysozyme C and leucine-rich alpha-2 glycoprotein (related to vascular damage and heart failure). High leucine-rich alpha-2 glycoprotein associated with higher mortality in stage 5 CKD patients on HD. Comparison: ESRD (stage 5 on HD) vs. earlier CKD stages (2-3). Sample type: Plasma. Platform: LC-MS/MS.

##### 2.2.6 Gene Co-expression Network Construction

Data Source and Rationale

Although numerous studies have performed co-expression analyses related to kidney disease, most focus on linking gene modules to external traits rather than providing summary statistics for all gene-gene relationships. Because such summary statistics were not publicly available, we conducted a de novo analysis to generate a resource of kidney-tissue-specific co-expression relationships.

Weighted gene co-expression network analysis (WGCNA) [39] was performed using bulk RNA-seq data from the Kidney Precision Medicine Project (KPMP, https://www.kpmp.org/), comprising samples from patients with acute kidney injury (AKI) and chronic kidney disease (CKD). All analyses were conducted using the R package WGCNA [39]. This analysis provides an initial resource of kidney-tissue-specific co-expression relationships for integration into the KD Atlas.

Sample Selection and Quality Control

The initial dataset comprised 31 samples. Quality control steps were as follows:

- Outlier detection: hierarchical clustering and PCA identified 5 outlier samples, which were excluded.
- Gene filtering: genes with low expression across samples were removed according to WGCNA guidelines.

The final dataset contained 26 samples for network construction.

WGCNA Pipeline

- **Variance stabilization**: DESeq2 variance-stabilizing transformation (VST) was applied.
- **Soft-thresholding**: the pickSoftThreshold function identified an appropriate power ($\beta = 18$) to approximate scale-free topology.
- **Adjacency and TOM**: Pearson correlation matrices were transformed into a signed adjacency matrix and then into a topological overlap matrix (TOM) to quantify gene-gene interconnectedness.
- **Module detection:** hierarchical clustering of 1-TOM followed by dynamicTreeCut identified modules of co-expressed genes (minimum module size = 30).
- **Module merging**: modules with highly similar eigengenes (height $< 0.25$) were merged, resulting in 17 distinct co-expression modules.

- **Network export:** gene pairs in each module, along with TOM values and module annotations, were exported using exportNetworkToCytoscape() for subsequent statistical assessment.

Statistical Filtering and Edge Selection

Gene-gene correlations were computed for all module gene pairs. P-values were calculated using corPvalueStudent() and adjusted with Bonferroni correction. Only positively correlated gene pairs with adjusted p-value < 0.05 were retained.

Integration into KD Atlas

The filtered co-expression data obtained after statistical assessment were integrated into the KD Atlas.

### 3. Network Construction and Abstraction

#### 3.1 Phase 1: Detailed Network Construction

Following the step-wise multi-omics integration strategy described in Section 1.2 , all kidney disease datasets were collected, preprocessed and integrated into a detailed, multi-layered network structure that preserves study- and dataset-specific information. The network was implemented in Neo4j (version 4.4.3) [40] to enable efficient querying and exploration of complex biological relationships.

Nodes represent molecular and phenotypic entities, including genes, transcripts, proteins, metabolites, SNPs, and traits, with additional source nodes corresponding to individual studies. Data-driven statistical associations between entities were encoded via intermediate nodes containing summary statistics (e.g., effect sizes, p-values) and linked to the originating study nodes to preserve provenance. The network includes the three categories of relationships defined in Section 1.3: intra-omics, inter-omics and phenotype-specific links.

#### 3.2 Phase 2: Network Abstraction and Simplification

The detailed network was projected into a simplified, abstracted view optimized for user interaction, following procedures described previously [1]. Key steps included:

- **SNP-to-gene projection:** Genetic variants were assigned to genes based on genomic location (gene bodies, flanking regions and regulatory elements from ENCODE and FANTOM5 [41]) and functional evidence from tissue-specific eQTL and pQTL data (GTEx v8 [5]). One-to-many SNP-to-gene mappings were retained,

and gene-wise significance thresholds were adjusted for the number of mapped SNPs.
- **Transcript and protein projection**: Transcripts were mapped to genes using Ensembl annotations, and proteins were linked to genes via transcript mappings, enabling aggregation of transcriptomic and proteomic information at the gene level.
- **Metabolite consolidation**: Metabolites measured across multiple platforms were consolidated into meta-metabolite nodes through manual curation. For example, metabolites on the Metabolon platform sharing biochemical names and chemical identifiers but differing COMP IDs were merged, reducing redundancy while retaining platform-specific annotations.

The abstracted network contains four primary node types:

- Genes: Protein-coding genes
- Metabolites (metMeta): Consolidated metabolites
- Traits: Individual kidney disease phenotypes and biomarkers
- Meta-traits: Collections of related phenotypes

Edge summarization and significance filtering: Associations between molecular entities were aggregated at the gene or metabolite level, while all underlying variant- and protein-level information was maintained in detailed edge annotations. Filtering of edges was performed using study-specific significance thresholds (**Table S6**) or thresholds selected by the user for genetic associations, either genome-wide ($p \leq 5\times10^{-8}$) or gene-wise. Gene-wise thresholds were defined as $p \leq 0.05/n$, where n represents the number of SNPs mapped to a given gene.

### 4. Web Interface Implementation and Technical Details

The KD Atlas is accessible via an interactive web interface that enables exploration of multi-layered omics data in the context of kidney disease phenotypes. Technical details of the implementation, including software versions and key functionalities, are provided below.

**Software and Environment**

- R version: 4.3.1
- R-Python integration: reticulate 1.34 with official Neo4j Python driver
- Deployment: ShinyProxy 2.3.1
- Interactive visualization: VisNetwork 2.0.9; 3D networks via threejs 0.3.3; network layouts via igraph 1.5.0 and qgraph 1.9.8

- UI enhancements: shiny 1.7.5.1, shinyWidgets 0.8.0, shinyjs 2.1.0, shinydashboard 0.7.2, shinydashboardPlus 0.7.0, shinybusy 0.3.1, scroller 0.1.1, shinycustomloader 0.9.0
- Data tables and plotting: DT 0.30, plotly 4.10.3, HiveR 0.3.63
- Data processing and manipulation: dplyr 1.1.3, purrr 1.0.2, stringr 1.5.0, mnormt 2.1.1, jsonlite 1.8.8

**Network Construction and Visualization**

- Graph Database: Neo4j 4.4.3 [40] stores nodes (genes, transcripts, proteins, metabolites, SNPs, traits, meta-traits) and edges representing statistically inferred relationships.
- Node and edge processing: Node degrees computed via igraph; network layouts precalculated using layout_with_drl, layout_with_graphopt, or qgraph.layout.fruchtermanreingold depending on user selection.
- Interactive features: Users can inspect edges to access detailed information stored in edge attributes. 2D and 3D network visualizations are rendered using VisNetwork and threejs; hive plots with HiveR; interactive bar/donut plots with plotly.

**Enrichment Analyses**

- Gene-level enrichment: Performed using topGO (v2.54.0) [42] for Gene Ontology terms, enrichR (v3.2) [43] for pathway and drug perturbation signatures, and gprofiler2 (v0.2.2) [44] for additional gene set analyses.
- Metabolite pathway enrichment: Conducted using Fisher's exact tests with platform-specific pathway annotations (Metabolon and Biocrates), applying false discovery rate correction.
- Differential expression enrichment: Genes were stratified into up-regulated, down-regulated, or combined sets, and classical pathway enrichment analyses were applied to each group.

### 5. KD Atlas User Interface Features

**Network Browser**

The network browser allows users to generate and visually explore kidney disease-related molecular subnetworks across multiple biological scales (**Figure S2**). It supports diverse query modalities centered on traits, genes or metabolites, enabling systematic investigation of molecular relationships underlying kidney disease.

Users can specify entities of interest through manual input or file upload. Query entities are annotated with multi-omics associations integrated within the KD Atlas resource. The initial query set can be expanded to include functional neighborhoods, filtered by biological context, and visualized as interactive networks.

Generated subnetworks are displayed as interactive graphs. Nodes, representing biological entities, can be clicked to access detailed information, while edges, representing associations, reveal additional details such as underlying SNPs and corresponding p-values.

Subnetworks can be downloaded as interactive HTML files, edge lists in CSV format, or in formats compatible with network analysis software such as Cytoscape. The following sections provide detailed explanations of the network browser features and parameters.

**Molecular Subnetwork Generation**

Users can provide one or more entities to query the KD Atlas resource. Eight distinct entry points are provided, organized into three primary categories (**Figure S2, layer 1**):

- **Trait- or meta-trait-centric subnetworks**: Users can query individual kidney disease-associated phenotypes (Trait) or collections of related phenotypes (Meta-trait) for trait-focused analyses.
- **Gene-centric subnetworks:** Gene-focused queries accept two identifier formats: Ensembl gene identifiers (Gene - Ensembl) or gene symbols (Gene - symbol). Both formats provide equivalent functionality and enable exploration of gene-trait associations, gene-metabolite relationships and gene-level molecular networks.
- **Metabolite- or pathway-centric subnetworks:** Metabolite-centered queries offer four complementary approaches: individual metabolites (Metabolite) identified by biochemical names, metabolites organized by broad metabolic categories (Metabolite - SuperPathway), metabolites grouped by specific biochemical processes (Metabolite - SubPathway) or metabolites classified by chemical structure (Metabolite - Classification).

**Network Expansion**

Prior to building the network, the initial set of input entities can be expanded to include 1-step or 2-step functional neighbors (**Figure S2, layer 2**). The available expansion methods depend on the query modality.

Gene-Centric Networks

For gene-focused queries (Gene – Ensembl or Gene – Symbol), functional neighbors can be defined using four types of molecular relationships. Users can select one or more expansion methods:

- **Coregulation (GTEx)**: Tissue-specific eQTL co-regulation networks from the Genotype-Tissue Expression Project, identifying genes sharing common regulatory variants
- **Co-expression:** Genes exhibiting similar expression patterns across samples.
- **Co-abundance:** Protein co-abundance relationships reflecting coordinated protein-level regulation
- **Protein-protein interaction:** Experimentally validated physical interactions between proteins (represented as protein-coding genes in the network)

Metabolite-Centric Networks

For metabolite-focused queries (Metabolite, Metabolite – SuperPathway, Metabolite – SubPathway, or Metabolite – Classification), expansion is performed using:

- **Gaussian Graphical Models (GGM):** Identify metabolites with direct statistical relationships.

Expansion Parameters

Users can enable or disable network expansion and specify the distance from the initial query (1-step or 2-step). Context filtering, if applied (see below), ensures that selected neighbors are relevant to the tissue or biological context of interest.

Once the input node set has been expanded, all nodes are annotated with associated entities from other omics layers, enabling the construction of fully integrated multi-omics networks.

**Significance Thresholds**

Associations between and within omics layers are filtered by applying significance thresholds (**Figure S2, layer 3**). For genetic associations—links between genes and metabolites (mGWAS) and genes and traits (GWAS)—users can select either gene-wise or genome-wide significance thresholds. The gene-wise threshold is defined as $p \leq 0.05/n$, where n is the number of SNPs annotated to a given gene using SNiPA. This Bonferroni-adapted correction is less stringent than the genome-wide threshold ($p \leq 5\times10^{-8}$) and typically yields more associations.

##### Context and Tissue-Specific Filtering

Edges can be filtered by sample type or tissue to provide context-specific, biologically relevant results (**Figure S2, layer 4**). The browser offers multiple filtering dimensions, which can be applied individually or in combination.

**Tissue filtering**: Links established through tissue-specific eQTL analysis (coregulation edges) derived from GTEx can be filtered by 49 tissues, including kidney cortex. Gene co-expression networks (co-expression edges) can also be filtered by tissue type, enabling selection of kidney-relevant contexts.

**Sample type filtering:** Genetic associations (gene-metabolite links from mQTL), metabolic associations, co-abundance relationships and partial correlation (GGM) edges can be filtered by sample type, which currently includes  plasma, serum and urine. This filtering enables users to focus on associations detected in biologically relevant sample contexts.

##### Interactive Visualization

The resulting molecular networks are rendered as interactive graphs where nodes represent biological entities (genes, metabolites and traits) and edges indicate evidence-supported associations (**Figure S2, layer 5**). The visualization interface provides multiple features to facilitate network exploration.

Users can inspect individual nodes to view detailed information including gene names (symbols, Ensembl IDs, chromosomal locations), metabolite annotations, or trait descriptions. Clicking on edges displays association details including the underlying SNPs and their corresponding p-values, allowing assessment of association strength and genetic architecture. The network graph supports various layout algorithms (2D, 3D, hive plots) to accommodate different network topologies and user preferences.

**Additional visualization options:** To assess the extent and direction of molecular dysregulation in kidney disease, differential analysis data can be projected onto the multi-omics networks. This includes genes that are differentially expressed – termed differentially expressed genes (DEGs) – and proteins that are differentially abundant – termed differentially abundant proteins (DEPs). Up-regulation, down-regulation or non-regulation at the transcriptional level is indicated by coloring gene nodes: red for upregulation, blue for downregulation, and muted sage green for unchanged expression. Differential protein abundance is indicated by coloring the borders of gene nodes using the same color scheme. DEG information is available for multiple tissue contexts, including blood, glomeruli, renal cortex and renal medulla.

#### Subnetwork Annotation and Analysis

##### Node Annotations

Entities within the generated molecular networks are annotated with comprehensive information accessible through interactive, searchable tables (**Figure S2, layer 6**). Metabolites are annotated with HMDB identifiers, biochemical names, super-pathways, sub-pathways and chemical classifications. Genes are annotated with chromosomal locations, Ensembl identifiers, and links to external databases, including GeneCards, Agora and Pharos. Kidney disease traits are provided with detailed phenotype descriptions. All biological entities are directly searchable within the network interface, allowing efficient identification and inspection of nodes even in large networks. Metabolite, gene and trait annotations can be downloaded as CSV files for downstream analysis.

##### Edge Annotations

Network edges are annotated with association-specific information accessible through interactive tables. Edge annotations include the connected biological entities (source and target nodes), minimal p-values representing the strongest statistical evidence for each association, edge types indicating the nature of the molecular relationship (e.g., GENETIC_TRAIT_ASSOCIATION, COEXPRESSION, METABOLIC_ASSOCIATION, COREGULATION, COABUNDANCE, PARTIAL_CORRELATION, and PROTEIN_INTERACTION), and tissue context where applicable.

##### Network Analysis

General network statistics are provided to aid quantitative network analysis and exploration **(Figure S2, layer 6**). Network summaries include total counts of genes, metabolites and traits, along with breakdowns of entities with trait associations. For genes, differential expression status is dynamically summarized across available tissue types (blood, glomeruli, renal cortex, renal medulla), with counts of upregulated, downregulated and unchanged genes in each context. Differential protein abundance is summarized for available DEP studies.

Network topology is characterized through node degree distributions, which can be visualized by node type (genes, metabolites and traits). Node type and edge type proportions are displayed as interactive donut charts, providing immediate visual insight into network composition.

##### Enrichment Analysis

Enrichment-based analysis can be performed for the generated molecular subnetworks using gene-level or metabolite-level annotations (**Figure S2, layer 6**). The biological

entities within the displayed network are tested for term enrichment against all entities within the KD Atlas resource. Results of enrichment analyses can be downloaded as CSV files.
**Gene-level enrichment employs multiple tools:** topGO (Gene Ontology enrichment for biological process, molecular function or cellular component), EnrichR (access to diverse gene set libraries), g:Profiler (functional enrichment across GO, KEGG, Reactome and disease annotations). Additionally, DEG enrichment tests for overrepresentation of differentially expressed genes within the subnetwork for user-specified tissue contexts, and DEP enrichment tests for overrepresentation of differentially abundant proteins. **Metabolite-level enrichment** utilizes Metabolon and Biocrates platform classifications, testing for overrepresentation of SuperPathways (broad metabolic categories), SubPathways (specific metabolic processes), or Classifications (chemical structure-based groupings).

**Data Export**

Networks and analytical results can be exported in multiple formats for downstream analysis and sharing (**Figure S2, layer 7**).

Interactive networks can be saved as standalone HTML files (.html) for presentation and sharing, allowing recipients to explore networks without accessing the KD Atlas platform. Edge lists, node annotations and enrichment results can be downloaded as comma-separated values (.csv) files.

For Cytoscape integration: Network data are formatted for compatibility with Cytoscape. Export options include Cytoscape graph (.json), Cytoscape style (.xml), and documentation (.pdf) providing guidance for importing and visualizing KD Atlas networks in Cytoscape.

### 6. Data Availability

- CKDGen Consortium: https://ckdgen.imbi.uni-freiburg.de/
- Genetic Epidemiology Group, University of Regensburg (source for Winkler et al. and Gorski et al. summary statistics): https://www.uni-regensburg.de/medizin/epidemiologie-praeventivmedizin/genetische-epidemiologie/gwas-summary-statistics/index.html
- KPMP: https://www.kpmp.org/
- Nephroseq: https://nephroseq.org/
- STRING-DB: https://string-db.org/
- HIPPIE: http://cbdm-01.zdv.uni-mainz.de/~mschaefer/hippie/
- IID: http://iid.ophid.utoronto.ca/

### SUPPLEMENTARY TABLES and FIGURES

**Table S1.** Summary of node and edge composition in the KD Atlas (abstracted).

| KD Atlas component | Count(n) |
|---|---|
| **Nodes** | |
| metaTrait (mt) | 6 |
| Trait (t) | 40 |
| gene (protein-coding) g | 20456 |
| DEG (>= 1 kidney disease phenotype) | 18983 |
| DEP (>= 1 kidney disease phenotype) | 1966 |
| metMeta (m) | 1375 |
| **Relationships** | |
| (t) - [ :PART_OF ] -> (mt) | 42 |
| (g) - [ :GENETIC_ASSOCIATION ] –> (t) | 34193 |
| (g) - [ :GENETIC_ASSOCIATION ] – >(m) | 98253 |
| (m) - [ :METABOLIC_ASSOCIATION ] –> (t) | 121 |
| (g) - [ :COEXPRESSION ] – (g) | 600562 |
| (g) - [ :PROTEIN_INTERACTION ] – (g) | 334416 |
| (g) - [ :PROTEIN_INTERACTION {confidence_level:High} ] – > (g) | 107924 |
| (g) - [ :PROTEIN_INTERACTION {confidence_level:Low} ] – > (g) | 61374 |
| (g) - [ :PROTEIN_INTERACTION {confidence_level:Medium} ] – > (g) | 165118 |
| (g) - [ :COREGULATION ] – > (g) | 449862 |
| (g) - [ :COREGULATION_{tissue} ] – > (g) | 4008088 |
| (g) - [ :COREGULATION_KIDNEY_CORTEX ] – > (g) | 6996 |
| (g) - [ :COABUNDANCE ] – (g) | 32406 |
| (m) - [ :PARTIAL_CORRELATION ] – (m) | 794 |

**Table S2.** Overview of Meta Traits and Corresponding Kidney Disease Traits.

| Meta trait | Trait |
|---|---|
| eGFR | eGFRcrea (overall)<br>eGFRcys (overall)<br>eGFRdecline<br>Rapid3<br>eGFR-decline (overall_adjBL)<br>eGFRcrea decline (Rapid3)<br>eGFRcrea<br>eGFRcys<br>creatinine-based eGFR(eGFRcrea)<br>cystatin C-based eGFR (eGFRcys)<br>eGFRcreat<br>eGFR creatinine<br>eGFR cystatin C<br>eGFRSCr<br>eGFRSCysC<br>annual eGFR change |
| CKD | chronic kidney disease (CKD)<br>Chronic Kidney Disease (CKD)<br>CKD<br>CKDi<br>CKDi25<br>eGFRcrea decline (CKDi25)<br>eGFR decline (CKD) |
| No CKD | eGFRdecline (noCKD)<br>Rapid3 (noCKD) |
| | eGFR decline (Diabetes)<br>eGFR-decline_overall (Diabetes) |

| | |
|---|---|
| Diabetes | eGFRcrea (Diabetes)<br>eGFR_DM (Diabetes)<br>UACR (Diabetes) |
| No Diabetes | eGFRcrea (no Diabetes)<br>eGFR_noDM (no Diabetes)<br>UACR (no Diabetes) |
| UACR | UACR<br>UACR overall |
| Others | Microalbuminuria (MA)<br>Urate<br>blood urea nitrogen(BUN)<br>incident kidney failure<br>KF+AKI |

**Table S3.** Kidney Disease Traits Integrated into the KD Atlas and Their Sources.

| Trait | Description | Study /Source | Type |
|---|---|---|---|
| eGFR decline (CKD) | Annual change in eGFR among participants with CKD at baseline, representing genetic effects on CKD progression | Gorski et al., 2022 | GWAS |
| eGFR decline (Diabetes) | Annual change in eGFR among participants with diabetes at baseline, adjusted for age and sex, reflects genetic effects on kidney function decline in diabetes. | Gorski et al., 2022 | GWAS |
| eGFR-decline (overall_adjBL) | Annual change in eGFR across all participants adjusted for age, sex, and baseline eGFR. Adjustment for baseline eGFR may induce collider bias for variants associated with baseline eGFR. | Gorski et al., 2022 | GWAS |
| eGFR-decline_overall (Diabetes) | Annual change in eGFR across all participants adjusted for age, sex, and diabetes status. | Gorski et al., 2022 | GWAS |

| | | | |
|---|---|---|---|
| eGFR_DM (Diabetes) | Estimated GFR among individuals with diabetes; used to identify genetic loci influencing kidney function specifically in diabetic participants. | Winkler et al., 2022 | GWAS |
| eGFR_noDM (no Diabetes) | Estimated GFR among individuals without diabetes; used to identify genetic loci influencing kidney function independently of diabetes status. | Winkler et al., 2022 | GWAS |
| eGFRcrea decline (CKDi25) | Binary phenotype defining rapid kidney function decline as ≥25% eGFRcrea loss during follow-up, with movement from eGFRcrea ≥60 mL/min/1.73$m^2$ at baseline to <60 mL/min/1.73$m^2$ at follow-up, compared to controls maintaining eGFRcrea ≥60; adjusted for age, sex, and baseline eGFR. | Gorski et al., 2021 | GWAS |
| eGFRcrea decline (Rapid3) | Binary phenotype defining rapid kidney function decline as annual eGFRcrea decrease >3 mL/min/1.73$m^2$, compared to “no decline” (1 to +1 mL/min/1.73$m^2$ per year); adjusted for age, sex, and baseline eGFR. | Gorski et al., 2021 | GWAS |
| creatinine-based eGFR(eGFRcrea) | eGFR based on serum creatinine among European ancestry participants. | Stanzick et al., 2021 | GWAS |
| cystatin C-based eGFR (eGFRcys) | eGFR based on serum cystatin C among European ancestry participants. | Stanzick et al., 2021 | GWAS |
| Urate | Serum urate concentration among individuals of European-American ancestry. | Tin et al., 2019 | GWAS |
| eGFRcreat | Estimated glomerular filtration rate based on creatinine, analyzed in individuals of European ancestry. | Wuttke et al., 2019 | GWAS |
| blood urea nitrogen(BUN) | Blood urea nitrogen among individuals of European ancestry, analyzed as an alternative marker of kidney function to complement eGFR-based measures. | Wuttke et al., 2019 | GWAS |
| CKD | CKD status among individuals of | Wuttke et al., 2019 | GWAS |

| | | | |
|---|---|---|---|
| | European ancestry, analyzed across ≥23 studies to detect loci associated with clinically diagnosed CKD. | | |
| UACR | Urinary albumin-to-creatinine ratio. The ratio of urinary albumin (a protein) to creatinine concentration in a spot urine sample. It serves as a quantitative measure of kidney damage. | Teumer et al., 2019 | GWAS |
| eGFRcrea (overall) | eGFR based on serum creatinine across all participants of European ancestry. | Gorski et al., 2017 | GWAS |
| eGFRcys (overall) | eGFR based on serum cystatin C across all participants of European ancestry. | Gorski et al., 2017 | GWAS |
| eGFRcrea (Diabetes) | eGFR based on serum creatinine among individuals with diabetes, used to examine kidney function specifically in diabetic participants. | Pattaro et al., 2016 | GWAS |
| eGFRcrea | eGFR based on serum creatinine across all participants of European ancestry | Pattaro et al., 2016 | GWAS |
| eGFRcrea (no Diabetes) | eGFR based on serum creatinine among individuals without diabetes, assessing genetic effects on kidney function independent of diabetes status. | Pattaro et al., 2016 | GWAS |
| eGFRcys | eGFR based on serum cystatin C | Pattaro et al., 2016 | GWAS |
| chronic kidney disease (CKD) | Chronic kidney disease status based on reduced eGFR (<60 ml/min per 1.73 $m^2$) among participants of European ancestry. | Pattaro et al., 2016 | GWAS |
| UACR (Diabetes) | Urinary albumin-to-creatinine ratio among individuals with diabetes. | Teumer et al., 2015 | GWAS |
| UACR | Urinary albumin-to-creatinine ratio (mg/g) | Teumer et al., 2015 | GWAS |
| UACR (no Diabetes) | Urinary albumin-to-creatinine ratio among individuals without diabetes, assessing genetic effects on albuminuria independent of diabetes status. | Teumer et al., 2015 | GWAS |
| Microalbuminuria (MA) | Microalbuminuria: UACR >25 mg/g | Teumer et al., 2015 | GWAS |

| | | | |
|---|---|---|---|
| | (women), >17 mg/g (men) | | |
| eGFRdecline (CKD) | Annual change in eGFR among participants with baseline CKD (eGFR <60 ml/min per 1.73 m²). | Gorski et al., 2015 | GWAS |
| eGFRdecline | Annual change in estimated glomerular filtration rate (eGFR, ml/min per 1.73 m² per year) across all participants, where a positive value represents decline and a negative value represents increase. | Gorski et al., 2015 | GWAS |
| eGFRdecline (noCKD) | Annual change in eGFR among participants without CKD at baseline (eGFR ≥60 ml/min per 1.73 m²). | Gorski et al., 2015 | GWAS |
| Rapid3 | Individuals with the fastest eGFR decline across all participants, defined as annual eGFR loss ≥3 ml/min per 1.73 m². | Gorski et al., 2015 | GWAS |
| Rapid3 (noCKD) | Rapid kidney function decline (≥3 ml/min per 1.73 m² per year) restricted to participants without CKD at baseline. | Gorski et al., 2015 | GWAS |
| CKDi | Incident CKD phenotype: selects individuals free of CKD at baseline who develop CKD stage 3 or higher (eGFR <60 ml/min per 1.73 m²) during follow-up. | Gorski et al., 2015 | GWAS |
| CKDi25 | Incident CKD with ≥25% decline in eGFR from baseline to identify individuals reaching CKD stage 3 after a substantial kidney function decline. | Gorski et al., 2015 | GWAS |
| eGFR creatinine | eGFR based on serum creatinine | Peggy et al., 2016 | MWAS |
| eGFR cystatin C | eGFR based on cystatin C | Peggy et al., 2016 | MWAS |
| Chronic Kidney Disease (CKD) | CKD defined as eGFRcrea <60 ml/min/1.73 m². | Peggy et al., 2016 | MWAS |
| eGFRSCr | Estimated glomerular filtration rate based on standardized serum creatinine levels (CKD-EPI equation) | Goek et al., 2012 | MWAS |

| | | | |
|---|---|---|---|
| eGFRSCysC | Estimated glomerular filtration rate based on serum cystatin C levels (CKD-EPI equation). | Goek et al., 2012 | MWAS |
| incident kidney failure | Time to kidney failure (KF), defined as a composite of initiation of dialysis, kidney transplantation or death due to foregoing dialysis. This endpoint represents progression to end-stage kidney disease requiring kidney replacement therapy or its clinical equivalent. | Steinbrenner et al., 2021 | MWAS |
| KF+AKI | Combined endpoint of kidney failure and acute kidney injury (AKI), where AKI was defined according to Acute Kidney Injury Network criteria stage 3 per the 2012 KDIGO guideline, including temporary dialysis. This composite outcome captures severe deterioration in kidney function due to either sustained or acute events. | Steinbrenner et al., 2021 | MWAS |
| annual eGFR change | Annual change in estimated glomerular filtration rate (eGFR) calculated as the difference between eGFR at baseline (S4) and follow-up (F4), divided by the number of years between visits. | Goek et al., 2012 | MWAS |

**Table S4.** Data Categories in the KD Atlas: Retained, Replaced, and Newly Added.

| Data Category | AD Atlas [1] | KD Atlas | Status | Notes |
|---|---|---|---|---|
| **Knowledge Databases** | | | | |
| Gene–transcript–protein mappings | Ensembl v97 | Ensembl v97 | Retained | Disease-agnostic |
| SNP annotations | SNiPA v3.3 | SNiPA v3.3 | Retained | Disease-agnostic |
| Tissue-specific eQTLs | GTEx v8 (49 tissues) | GTEx v8 (49 tissues) | Retained | Disease-agnostic |
| **Population-Based Data** | | | | |
| Metabolite mGWAS | Suhre et al. 2011; Shin et al. 2014; Raffler et al. 2015; Draisma et al. 2015; | Retained from AD Atlas | Retained | Disease-agnostic |

| | | | | |
|---|---|---|---|---|
| | Long et al. 2017 | | | |
| Protein partial correlations | Suhre et al. 2017 | Retained from AD Atlas | Retained | Disease-agnostic |
| Metabolite partial correlations | Krumsiek et al. 2012 | Retained from AD Atlas | Retained | Disease-agnostic |
| **Disease-Specific Data** | | | | |
| AD datasets (GWAS, MWAS, DE, co-expression) | Present | – | Replaced | Replaced with kidney-specific equivalents |
| Kidney disease datasets (GWAS, MWAS, DE, co-expression) | – | CKDGen consortium, Nephroseq, KORA, GCKD | Replaced | Kidney-specific integration |
| **Newly Added Data** | | | | |
| Plasma–urine metabolite mQTLs | – | Schlosser et al. 2023 | Added | Kidney-specific |
| New Protein partial correlations | – | Suhre et al. 2024 | Added | Extended protein co-abundance relationships |
| Protein–protein interaction Databases | – | STRING / HIPPIE / IID | Added | Broad coverage of interactions |

**Table S5.** Number of measured metabolites/compounds consolidated to one "meta-metabolite" in the KD Atlas. For example, n=395 meta metabolites consolidate 2 measured metabolites.

| **Number of measured metabolites/compounds consolidated to one "meta-metabolite"** | **n (number of meta-metabolites)** |
|---|---|
| 1 = no consolidation | 780 |
| 2 | 395 |
| 3 | 151 |
| 4 | 32 |

| | |
|---|---|
| 5 | 8 |
| 6 | 7 |
| 7 | 1 |
| 8 | 1 |

**Table S6.** Data sources and references.

| Data Type | Edge Type | Significance threshold | Cohort | Reference |
|---|---|---|---|---|
| **KNOWLEDGE DATABASES** | | | | |
| PPI | **PROTEIN_INTERACTION** | STRING ≥ 0.7; HIPPIE ≥ 0.73 High confidence: 3 databases<br>Medium confidence: 2 databases<br>Low confidence: 1 database | STRING v11.0<br>HIPPIE v2.0<br>IID v2021.05 | Szklarczyk et al. (2019) [11]<br>Alanis-Lobato et al. (2016) [12]<br>Kotlyar et al. (2016) [13] |
| **POPULATION-BASED DATA** | | | | |
| eQTL | **COREGULATION** | q-value ≤ 0.05 | GTEx v8 | GTEx Consortium (2020) [39] |
| mQTL | **GENETIC_ASSOCIATION** | P ≤ 0.0001[#] | KORA, TwinsUK | Suhre et al. (12) (2011) [7] |
| | | P ≤ 0.001[#] | KORA, TwinsUK | Shin et al. (2014) [3] |
| | | P ≤ 0.0001[#] | SHIP-0, KORA | Raffler et al. (2015) [4] |
| | | P ≤ 0.0001[#] | Meta-analysis | Draisma et al. (2015) [5] |
| | | P ≤ 0.0001[#] | TwinsUK | Long et al. (2017) [6] |
| | | Study-reported genome-wide significant associations | GCKD, ARIC, UK Biobank | Schlosser et al. (2023) [30] |
| GGM | **COABUNDANCE** | 0.05/all possible edges | KORA | Suhre et al. (2017) [9] |

| | | | | |
|---|---|---|---|---|
| | | $P < 4.7 \times 10^{-8}$ 0r \|pcor\| > $1.76 \times 10^{-3}$ | UK Biobank | Suhre et al. (2024) [10] |
| GGM | **PARTIAL_CORRELATION** | p-value ≤ 7.96e-7 and abs(cor) ≤ 0.1603 | KORA | Krumsiek et al. (2012) [8] |
| **KIDNEY-SPECIFIC GENOME-WIDE ASSOCIATION STUDIES (GWAS)** | | | | |
| traitQTL | **GENETIC_ASSOCIATION** | P ≤ 0.05[#] | Meta-analysis (CKDGen, UK Bio, MVP, MGI, HUN) | Winkler et al. (2022) [15] |
| | | P ≤ 0.05[#] | CKDGen & UK BB | Gorski et al. (2022) [16] |
| | | P ≤ 0.05[#] | Meta-analysis (CKDGen & UK Biobank)[*] | Stanzick et al. (2021) [17] |
| | | P ≤ 0.05[#] | Meta-analysis (CKDGen & UK Biobank)[*] | Gorski et al. (2021) [18] |
| | | P ≤ 0.05[#] | Meta-analysis | Tin et al. (2019) [19] |
| | | P ≤ 0.05[#] | Meta-analysis (CKDGen & UK Biobank)[*] | Teumer et al. (2019) [20] |
| | | P ≤ 0.05[#] | Metana-analysis (CKDGen, MVP) | Wuttke et al. (2019) [21] |
| | | P ≤ 0.05[#] | Metana-analysis (1000 Genomes imputed) | Gorski et al. (2017) [22] |
| | | P ≤ 0.05[#] | Metana-analysis (CKDGen) | Pattaro et al. (2016) [23] |
| | | P ≤ 0.05[#] | Metana-analysis | Teumer et al. (2015) [24] |
| | | P ≤ 0.05[#] | Metana-analysis | Gorski et al. (2015) [25] |
| **KIDNEY-SPECIFIC METABOLOME-WIDE ASSOCIATION STUDIES (MWAS)** | | | | |
| mWAS | **METABOLIC_ASSOCIATION** | p-value ≤ 0.05/(number of metabolites tested for that endpoint) | GCKD | Steinbrenner et al. (2021) [26] |
| | | $P < 1.0 \times 10^{-4}$ ( 0.05 / | KORA, TwinsUK, | Peggy et al. (2016) [27] |

| | | | |
|---|---|---|---|
| | 488) | AASK | |
| | P < 3.6 × $10^{-4}$ (0.05/140) | KORA | Goek et al. (2013) [28] |
| | P < 3.3 $10^{-4}$ (0.05/151) | KORA | Goek et al. (2012) [29] |
| **DIFFERENTIAL GENE EXPRESSION (DEG)** | | | |
| **DEG** | P < 0.01 & \|log2FC\| > 1 | Kidney cortex vs. medulla | Haug et al. (2024) [31] |
| | P < 0.05 | IgAN vs. healthy donors (blood monocytes) | Cox et al. (2015) [32] |
| | FDR ≤ 0.1 | FSGS/collapsing FSGS/MCD vs. control (glomeruli) | Hodgin et al. (2010) [33] |
| | P < 0.001 | Aging kidney (cortex and medulla) | Rodwell et al. (2004) [34] |
| **DIFFERENTIAL PROTEIN ABUNDANCE (DEP)** | | | |
| **DEP** | P < 0.05 | CKD stage 5 (hemodialysis) vs. stage 2-3 (plasma) | Glorieux et al. (2015) [35] |
| **GENE CO-EXPRESSION NETWORKS** | | | |
| **COEXPRESSION** | Bonferroni-adjusted P < 0.05 | KPMP: AKI and CKD kidney tissue | This study |

[*]The primary data for this meta-analysis came from the CKDGen Consortium and the UK Biobank cohort; [#]results up to reported threshold are integrated in resource and genome-wide or gene-wide significance is applied to generate context-specific networks

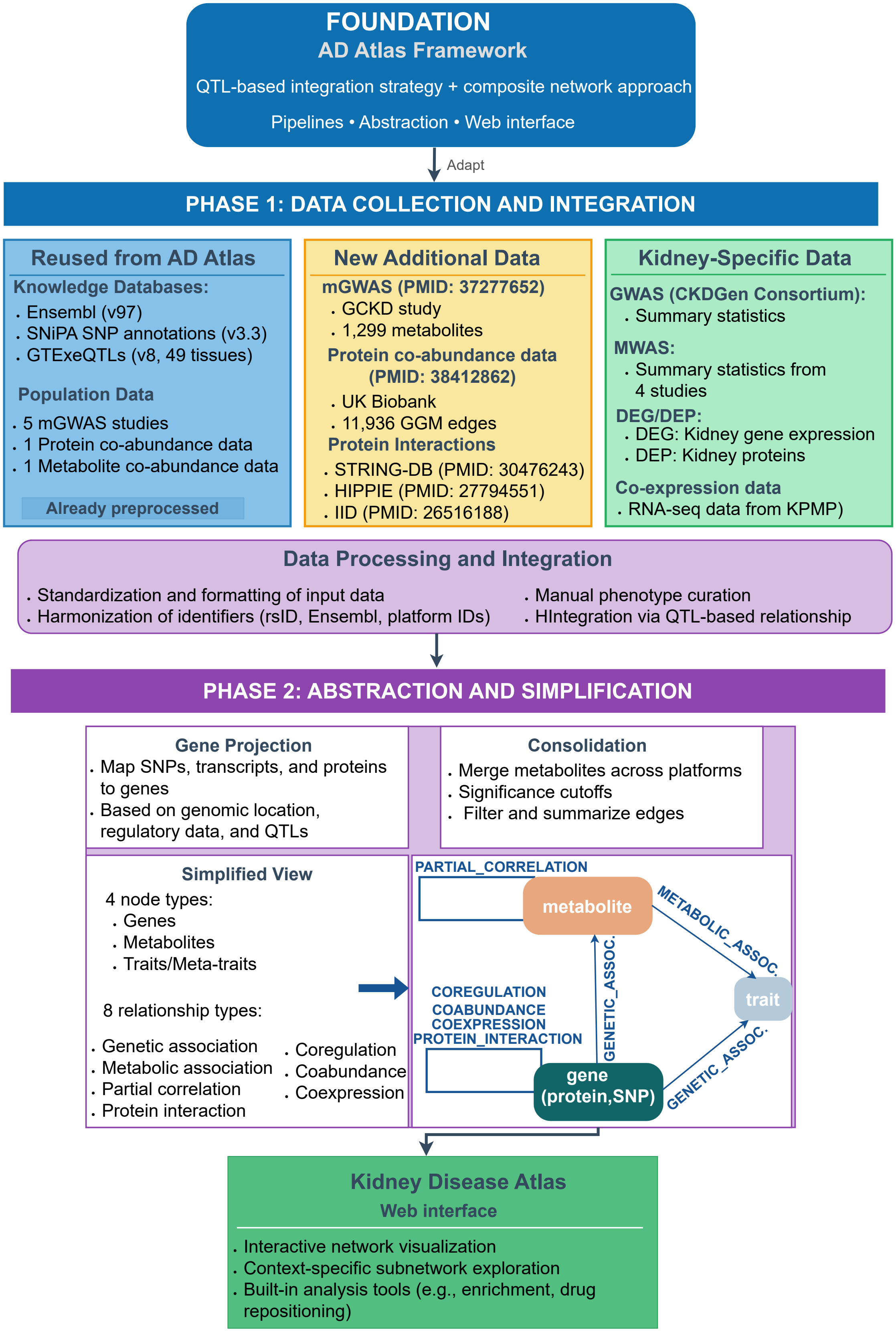
FOUNDATION
AD Atlas Framework
QTL-based integration strategy + composite network approach
Pipelines • Abstraction • Web interface
Adapt
PHASE 1: DATA COLLECTION AND INTEGRATION
Reused from AD Atlas
Knowledge Databases:
. Ensembl (v97)
. SNiPA SNP annotations (v3.3)
. GTExeQTLs (v8, 49 tissues)
Population Data
. 5 mGWAS studies
. 1 Protein co-abundance data
. 1 Metabolite co-abundance data
Already preprocessed
New Additional Data
mGWAS (PMID: 37277652)
. GCKD study
. 1,299 metabolites
Protein co-abundance data (PMID: 38412862)
. UK Biobank
. 11,936 GGM edges
Protein Interactions
. STRING-DB (PMID: 30476243)
. HIPPIE (PMID: 27794551)
. IID (PMID: 26516188)
Kidney-Specific Data
GWAS (CKDGen Consortium):
. Summary statistics
MWAS:
. Summary statistics from 4 studies
DEG/DEP:
. DEG: Kidney gene expression
. DEP: Kidney proteins
Co-expression data
. RNA-seq data from KPMP)
Data Processing and Integration
. Standardization and formatting of input data
. Harmonization of identifiers (rsID, Ensembl, platform IDs)
. Manual phenotype curation
. HIntegration via QTL-based relationship
PHASE 2: ABSTRACTION AND SIMPLIFICATION
Gene Projection
. Map SNPs, transcripts, and proteins to genes
. Based on genomic location, regulatory data, and QTLs
Consolidation
. Merge metabolites across platforms
. Significance cutoffs
. Filter and summarize edges
Simplified View
4 node types:
. Genes
. Metabolites
. Traits/Meta-traits
8 relationship types:
. Genetic association
. Metabolic association
. Partial correlation
. Protein interaction
. Coregulation
. Coabundance
. Coexpression
PARTIAL_CORRELATION
metabolite
METABOLIC_ASSOC.
trait
GENETIC_ASSOC.
COREGULATION
COABUNDANCE
COEXPRESSION
PROTEIN_INTERACTION
gene (protein,SNP)
GENETIC_ASSOC.
Kidney Disease Atlas
Web interface
. Interactive network visualization
. Context-specific subnetwork exploration
. Built-in analysis tools (e.g., enrichment, drug repositioning)

**Figure S1. Overview of the KD Atlas development and data integration pipeline.** The KD Atlas was built upon the AD Atlas [1] framework foundation, retaining its core QTL-based integration strategy, composite network approach, two-phase pipeline, and web interface. Phase 1 encompasses data collection and integration from three sources: disease-agnostic data reused from the AD Atlas (blue), new population-based datasets (yellow), and kidney disease-specific data (green), followed by preprocessing, formatting and integration into a comprehensive network. Phase 2 involves abstraction and simplification through gene projection, consolidation and filtering to create an intuitive view optimized for user interaction via the web interface.

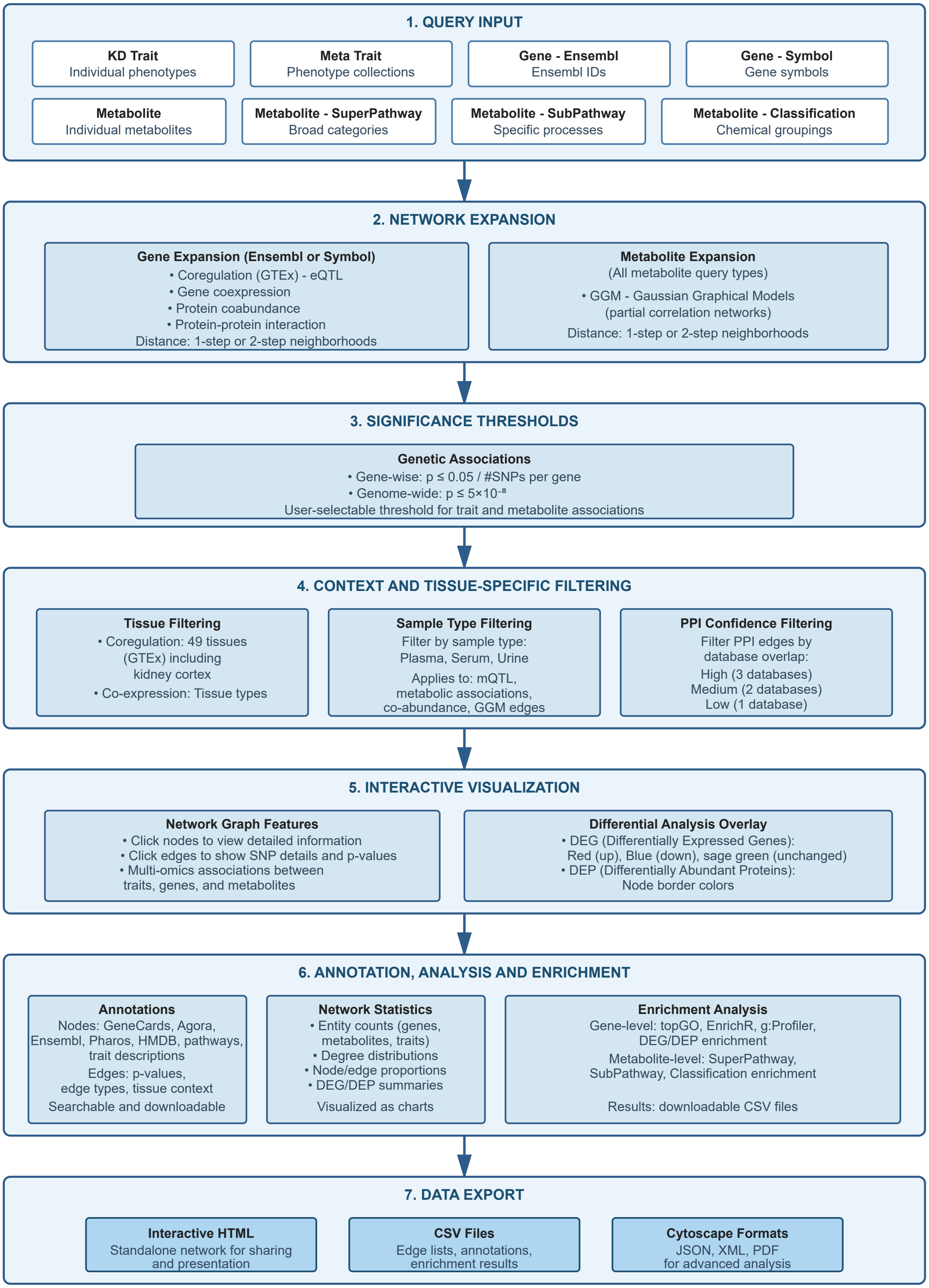

Manual input or file upload supported • Context filtering applied throughout • Multiple layouts (2D, 3D, hive plots)

**Figure S2. Detailed workflow of the KD Atlas network browser interface.** The network browser workflow consists of seven steps: (1) Query input via trait-centric, gene-centric, or metabolite-centric entry points; (2) Optional network expansion through functional relationships; (3) Application of significance thresholds for genetic associations; (4) Context and tissue-specific filtering; (5) Interactive visualization with differential expression overlay (DEG: Differentially Expressed Gene; DEP: Differentially Expressed Protein); (6) Network annotation, statistics, and enrichment analysis; (7) Data export in multiple formats including interactive HTML, CSV files and Cytoscape-compatible formats.
